# Beyond the medical file: a scoping review on patients’ perspectives on guideline-oriented depression treatment in primary care

**DOI:** 10.1101/2023.12.27.23297265

**Authors:** Katharina Biersack, Heribert Sattel, Petra Schönweger, Lea Kaspar, Nadine Lehnen, Jochen Gensichen, Peter Henningsen, the POKAL group

**Affiliations:** Department of Psychosomatic Medicine and Psychotherapy, University Hospital of Technical University of Munich, Germany; Institute of Medical Information Processing, Biometry, and Epidemiology – IBE, Chair of Public Health and Health Services Research, Munich, Germany; Pettenkofer School of Public Health, Munich, Germany; Institute of General Practice and Family Medicine, University Hospital of Ludwig-Maximilian-University Munich, Germany

**Keywords:** scoping review, clinical practice guideline, primary care, depression, patient perspective

## Abstract

**Objectives:** Depressive disorders are common in the primary care setting. Primary care practitioners must deal with different disorders and keep up with evidence-based treatment. Clinical practice guidelines (CPG) offer accessible information about up-to-date care but are poorly implemented. Research on the implementation of CPGs has focused on the practitioners’ perspective but has neglected the patients’ perspective largely.

This scoping review aimed to identify terms related to the concept of ‘patients’ perspectives on depression treatment in primary care building a comprehensive framework and to identify researched barriers and facilitators to partaking in care.

**Methods:** We conducted a scoping review on Medline and Psycinfo. Eligible publications contained information from the patients’ point of view on depression treatment in primary care in OECD member states. We used the PCC-framework to obtain inclusion criteria. Publications until August 2nd 2023 were considered.

**Results:** We included 232 publications. Current literature focuses on behavioral and easily collected measures like satisfaction and on patient-sided barriers and facilitators to adherence. Other treatment-related behaviors are less researched. Patients often report exclusively or mainly physical symptoms in their visits which can impede diagnosis.

**Conclusion:** This review provides a comprehensive framework for the concept. Research on the patients’ perspective on depression treatment in primary care is still inconclusive.

**Registration:** This review is registered via OSF (https://osf.io/p9rnc).

## 1. Introduction

Depressive disorders are very common in the community and often reason for disability and absenteeism [1,2]. Undetected and inadequately treated cases of depression are a burden to patients, health system, and society [3]. Most patients are not seen and treated in specialist care but at the office of their local GP [2]. Still, depression is often undiagnosed and, even after diagnosis, still untreated [4]. To this day, the reasons for these gaps are not completely clear, nor are there tailored and effective strategies.

Clinical practice guidelines (CPGs) have long been used to bridge the gap between best practice and real-world primary care. Most high and middle-income countries have national guidelines for depression treatment, developed by specialists and stakeholders [5]. However, many guidelines are not adequately implemented in primary care settings over long periods of time [6]. Despite their aim of improving accessibility to information and effective treatment strategies, CPGs face implementation challenges.

Primary care professionals deal with a wide range of disorders and must stay updated on numerous CPGs. As a result, not all guidelines are equally known and implemented. This can be attributed, in part, to guidelines themselves, as only a few provide effective implementation guidance. For example, the German national CPG for depression treatment mentions “implementation” only twice in its 257 pages with no clear guidance on the topic [7]. A systematic review by Lee et al. in 2020 concluded that so far guideline implementation is inadequately planned, reported, and measured globally [8].

On the practitioners’ side, barriers naturally emerge when striving for evidence-based treatment for all patients and disorders in primary care. Barriers to CPG usage in primary care, such as lack of education or limited consultation time, have been a subject of interest and research among clinicians and stakeholders [9]. A recent simulation study revealed that primary care professionals would require more than 26 hours per day to implement, use, and document according to all current CPGs. 1.6 hours would be solely dedicated to mood disorders [10].

To improve the implementation of evidence-based treatment for depressed patients in primary care, various reviews highlight systemic problems requiring large-scale policies [8,9,11]. However, it takes considerable time before these policies effectively enhance patient care. In the meantime, research should focus on identifying gaps and exploring alternative approaches to aid implementation.

One significant gap lies in understanding patients’ perspectives on treatment. Patient-reported and - relevant outcomes are a topic of growing interest as patient-centered care is required to decide whatis meaningful and valuable to the individual patient [12]. Patients’ perspectives, their values and preferences should be considered in the clinical decision-making process [13]. Up to now, patient-reported outcomes often consist in symptom-related questionnaires but do neglect patients’ point of view on domains such as satisfaction, expectations and contextual information [14]. Moreover, patient-relevant outcomes are underrepresented in the current literature despite being important in the treatment [15]. Currently, there are no frameworks or reviews available on how patients, as key stakeholders, perceive their care. Just recently, a review emphasized the need for increased patient participation in guideline development and to date only few guidelines involve patients in the process [5].

Could patients’ perspectives serve as the missing link between evidence and practice? Patients’ experiences with depression treatment in primary care, barriers to help-seeking, and factors affecting their understanding of and engagement in treatment are vital for effective implementation strategies. With this scoping review we provide a systematic overview to help guide future research.

## 2. Aim

This scoping review aimed at identifying concepts related to ‘patients’ perspectives on primary care’ in a systematic manner, to provide a comprehensive framework with domains and superordinate domains.

Primary aim was to identify the scientific methods and concepts related to patients’ perspectives on depression treatment in primary care.

Secondary we aimed to identify already researched barriers and facilitators towards evidence-based treatment of depression in primary care.

## 3. Methods

### 3.1. Search strategy

We conducted a scoping review searching the databases of Medline and Psycinfo with no limitation to publication dates up to August 2nd, 2023. By choosing these sources we felt confident to identify and screen all relevant publications to meet our research goals. We conducted our scoping review in accordance with the JBI Manual for Evidence Synthesis [16]. We registered the review and gave details of our method via Open Science Framework (https://osf.io/p9rnc).

### 3.2. Inclusion criteria

For the design of our review and the research questions we used the PCC-Framework (population, context, concept) recommended for scoping reviews by JBI which is applicable for quantitative, qualitative, and mixed-methods studies [16]. See table 1 for details. For the applied search terms see appendix 1. We included publications available in English, German, Spanish or French as we felt confident enough to judge and extract information in these languages.

**Table 1.** PCC-Framework.

|  |  |
| --- | --- |
| Population | Adult currently clinically depressed patients in OECD member states |
| Context | Primary care as defined by the authors, all available studies |
| Concept | Patients' perspectives on depression care i.e., their behavior, preconception, and experience |

#### 3.2.1. Population

We included studies with adult persons suffering from depression when being studied. We decided not to include studies solely on children and adolescents as treatment and needs are very specific and may be tailored more to age than to the disorder itself. We defined ‘depression’ as an, up-to-date, ICD or DSM based diagnosis before conducting the review.

We changed our approach to defining depression compared to our study protocol. It proved too exclusive to include only publications with a clear stated definition of the diagnosis of depression, as many publications included subjects diagnosed by their PCP without further specification. To tackle our goal to provide a scope to the field we decided to include all studies that stated how they diagnosed depression. We excluded several qualitative studies on the grounds that they refused to define depression because it contradicted their approach.

If the study included both depressed and dysthymic patients, it was still included for reasons of practicality. For the same reason, we included studies conducted in patients who were selected because of an antidepressant prescription if the authors stated that the prescription was for the treatment of depression and not for some other disorder e.g., anxiety. We also included publications in which the diagnosis was given by the PCP even if their approach was not clearly stated. We narrowed the search to OECD-member states for means of comparability of the studied context.

#### 3.2.2. Context

Because we were interested in the context of primary care i.e., how patients and other stakeholders are understanding and using it, we tried to be as inclusive as possible with the concept of primary care. We selected papers that referred to the setting as ‘primary care’ and defined the setting. Patients did not necessarily have to be recruited in primary care but had to give information about their diagnosis or treatment in that context. We included all ambulatory settings meeting our criteria.

#### 3.2.3. Concept

To define the concept of patients’ perspectives on care we scoped accessible and recent literature. We decided to include papers that contained information from the patients’ own account of their care and had to relate not just to symptomatology but also to the care itself. To find search terms beyond ‘perspective’ we looked for related topics which we balanced with existing Mesh-terms (See also registration). After finding subcategories for our main concept, we divided these into three conceptual domains: patients’ behavior, patients’ preconception, and patients’ experience.

##### 3.2.3.1. Preconception

We sought to include patients’ own concept of depression and treatment and its importance for the beginning and continuity of care. After scoping the literature, we identified various pre-existing concepts describing aspects of the mind related to action e.g., ‘attitude’, ‘belief’, ‘knowledge’, or ‘expectation’. To bring these different terms and concepts together, we agreed on the term ‘preconception’. The chosen term points to the idea of conditions preceding the patient-doctor-encounter we were interested in.

##### 3.2.3.2. Experience

To conceptualize terms describing the patients’ point of view while in treatment we used the term ‘Experience’. We included studies in which patients were a distinguishable group and their information added to the concept.

##### 3.2.3.3. Behavior

We decided to include behavioral means into our definition of the concept because behavior is directly related to motivation and can hint to potential barriers and facilitators. It is also easy to detect and describe from a research point of view. Among the terms we searched for were ‘consultation’ or ‘help seeking’. To look for patterns of symptom report, we included studies that used open ended measures to describe the complaints with which patients initially presented. We excluded studies in which symptoms were given via broad psychometric testing because these findings tend to be pre-empted by the questionnaires and are not indicative for spontaneous accounts. We used the search terms ‘symptoms’ and ‘complaints’ for that purpose.

### 3.3. Barriers, facilitators, and subpopulations

The second and third research question did not directly impact the search strategy or PCC-framework. By defining all above-mentioned concepts as inclusive as possible and as exclusive as needed we felt comfortable that we included all relevant publications to answer the remaining questions.

We defined ‘barriers and facilitators to depression treatment in primary care from the patients’ point of view’ as factors that impede or help with therapy-related patient behaviors. We sought to include all studies using the terms ‘barrier’ or ‘facilitator’ but did not want to exclude relevant papers researching outcomes falling under our definition which used different terms.

### 3.4. Review and synthesis process

We evaluated every step of the review process within our research group following a mixed deductive and inductive approach. After prescoping we decided on the superordinate domains i.e., ‘preconception’, ‘experience’, and ‘behavior’ which helped build the research strategy. After inclusion we extracted and grouped arising topics within domains and superordinate domains inductively.

We uploaded our searches of Medline and Psycinfo into the software of Rayyan and conducted the abstract screening via this platform. Screening was conducted by three independent raters continuously evaluating adherence to inclusion criteria, adapting them when needed.

## 4. Results

### 4.1. Search results

The conducted search of the databases identified 16.830 results. After the removal of 2771 duplicates, we included 14059 studies in the screening of title and abstract. Following the mentioned PCC-scheme we could exclude 13534 abstracts. Within the remaining 525 studies, 14 papers could not be retrieved even after directly contacting authors and libraries. During screening of full texts, we excluded 34 studies because they did not offer new data, 1 study because it lacked peer-review, 141 studies due to wrong population, 56 because of wrong context and 47 because of wrong concept. We included 232 studies into our data synthesis. The results are presented in a Preferred Reporting Items for Systematic Reviews and Meta-analyses for Scoping Reviews (PRISMA-ScR) flow diagram [17]. See figure 1.

**Figure 1.**
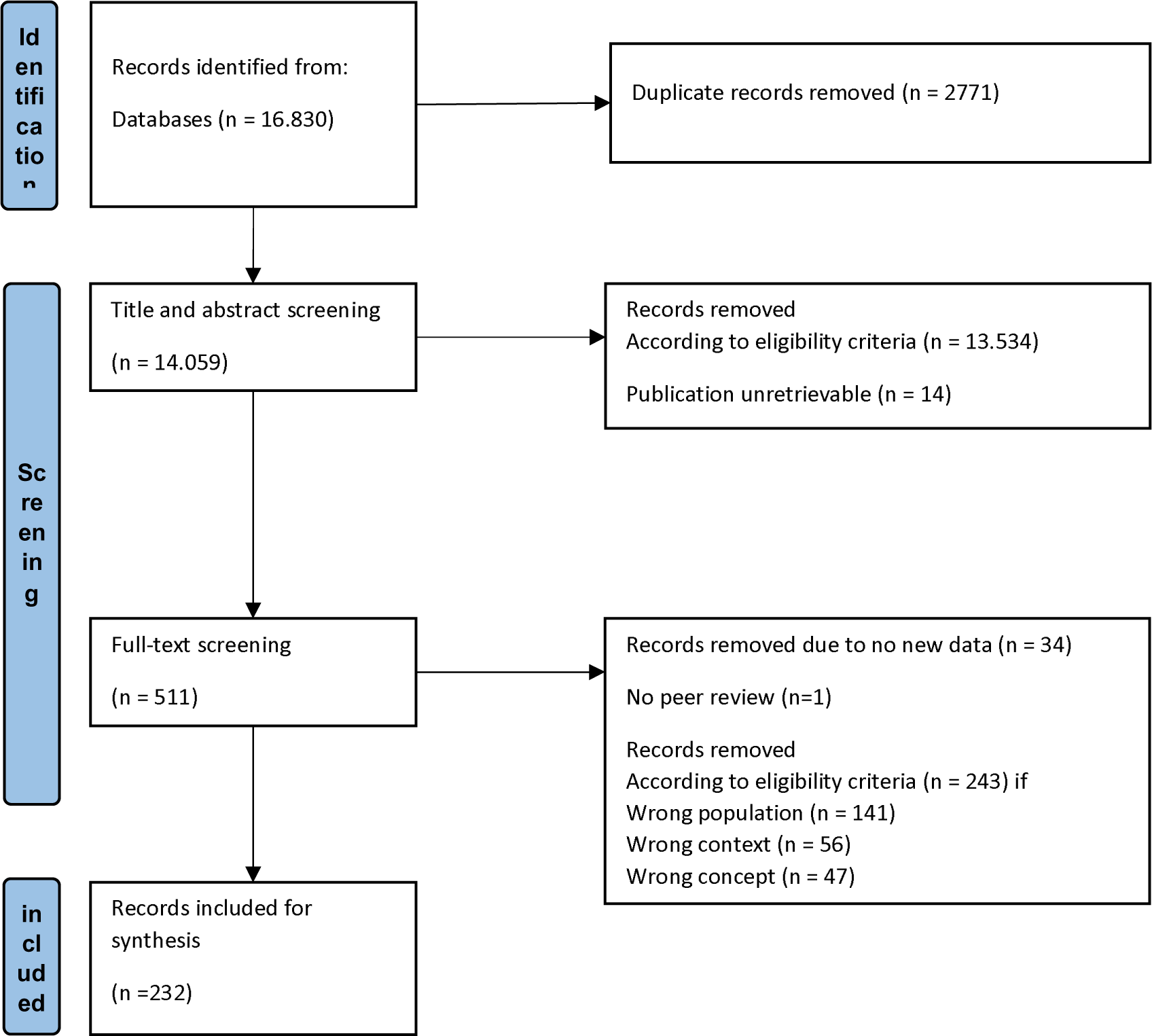
Flow diagram describing search strategy and results.

### 4.2. Studies’ characteristics and methodology

Within included articles, the grand majority (41%, n=95) were conducted in the US, 24% (n=54) in EU countries, 25% (n=57) in the UK and 6% (n=13) in Australia. The remaining studies were conducted in Canada (n=3), Israel (n=3), Japan (n=2), New Zealand (n=2) and Norway (n=1). Two studies included data from more than one location [18,19]. Because we excluded non-OECD states as context in our review showing the local distribution of included reports bears limited information.

Sixty three percent of included publications used exclusively quantitative measures for the looked-for outcomes, while 37% used a qualitative or mixed methods approach. For the distribution by year of publication see figure 2.

**Figure 2.**
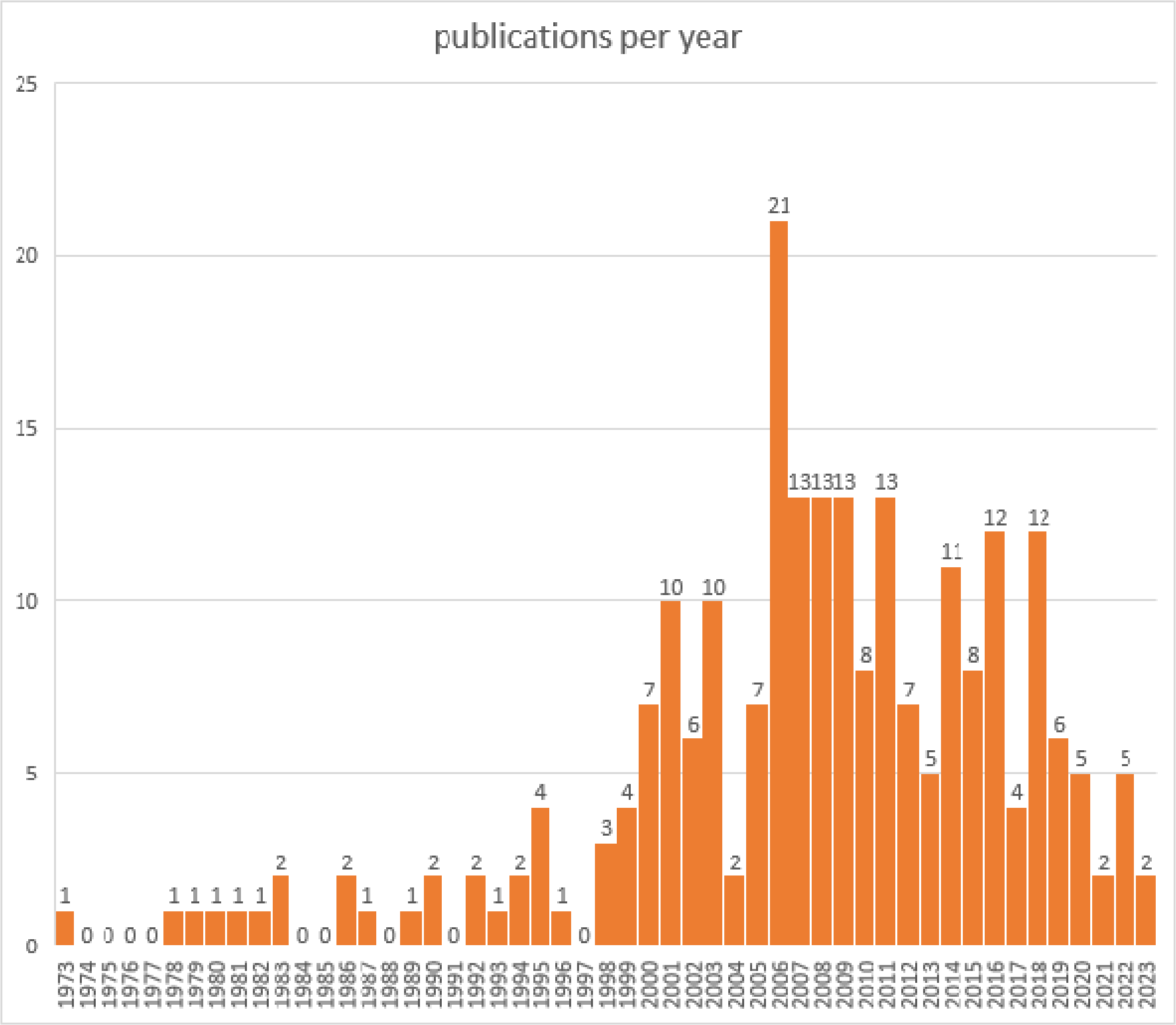
Temporal distribution of publications.

### 4.3. Population

Most publications (38,8%) used self-rating scales to define depression and identify subjects, most studies used the PHQ-9, PHQ-8, or PHQ-2. Twenty six percent defined ‘depressed patient’ as diagnosed by their primary care practitioner. Other publications used the prescription of antidepressants for depression, patient chart, or patient history, or gave details on used checklists for diagnosis. See also table 2.

**Table 2.** Description of depression diagnosis in included publications. Total numbers and percentages.

| Concept of depressed patient | Number of publications | % |
| --- | --- | --- |
| Diagnosis by PCP | 61 | 26,29 |
| Diagnostic interview | 47 | 20,26 |
| Self-rating | 46 | 19,83 |
| PHQ | 44 | 18,96 |
| Prescription of an antidepressant | 15 | 6,46 |
| Chart | 9 | 3,88 |
| Patient history | 7 | 3,02 |
| Checklist | 2 | 0,86 |
| Diagnosis by Author | 1 | 0,43 |

### 4.4. Context

We defined primary care as all outpatient settings dedicated to primary and generalist care. Table 3 gives details on used terms in the included publications. The most used name for the primary care setting was ‘general practice’ and ‘general practitioner’ (44%). Less used but trending in more recent publications was ‘collaborative care’ (2%).

**Table 3.** Description of primary care context in included publications. Total numbers and percentages.

| Concept of primary care | Number of publications | % |
| --- | --- | --- |
| General practice | 96 | 41,38 |
| Primary care clinic | 33 | 14,22 |
| Primary care practice | 19 | 8,19 |
| Health center | 11 | 4,74 |
| Family practice | 10 | 4,31 |
| Veteran care | 8 | 3,45 |
| Collaborative care | 1 | 0,43 |
| Aged-care facility | 1 | 0,43 |
| Family medicine clinic | 6 | 2,59 |
| Other | 10 | 4,31 |
| PC | 37 | 15,95 |

### 4.5. Patients’ perspectives on depression treatment in primary care

#### 4.5.1. Identified terms and domains

In an inductive process we identified relevant domains for the concept and the premeditated superordinate categories, i.e., preconception, experience, and behavior, served well as a framework for the established research terms. See table 4 for the grouped domains and superordinate domains as well as the number of their occurrences among the included publications.

**Table 4.** Patients’ perspectives on depression treatment in primary care: Number of occurrences of named topics, grouped into domains and superordinate domains within the premeditated framework i.e., ‘preconception’, ‘experience’ and ‘behavior’.

| Patients’ perspectives: | Number of publications on the | Sources |
| --- | --- | --- |

| Domain and topic | topic |  |
| --- | --- | --- |
| **Domain: Preconception** | **105** |  |
| 1. Attitude to care | 18 | [20–37] |
| 2. Concept of depression | 45 | [27,31,33,37–78] |
| 3. Stigma | 9 | [35,62,64,70,71,79–82] |
| 4. Entitlement to time | 1 | [83] |
| 5. Confidence | 2 | [22,84] |
| 6. Treatment expectations | 5 | [32,44,85–87] |
| 7. Knowledge | 3 | [71,81,88] |
| 8. Treatment preference | 22 | [24,60,70,73,86,89–105] |
| **Domain: Experience** | **128** |  |
| 1. Satisfaction | 18 | [31,68,88,99,101,106–118] |
| 2. Receiving information | 3 | [33,43,119] |
| 3. Reported relationship | 17 | [20,50,58,68,70,106,109,111,113,114,120–126] |
| 4. Acceptance of diagnosis and care | 9 | [32,68,71,127–132] |
| 5. Reported quality of care | 9 | [21,109,111,128,133–137] |
| 6. Experience of depression | 7 | [65,66,120,138–141] |
| 7. Experience of treatment | 41 | [31,32,38,40,41,44,58,63,67,68,70,85,90,101,112–115,119,129,130,134–136,142–158] |
| 8. Barriers to care | 14 | [37,41–43,64,121,157,159–165] |
| 9. Facilitators to care | 5 | [26,138,162,164,166] |
| 10. Participation in treatment decisions | 5 | [43,167–170] |
| **Domain: Behavior** | **150** |  |
| 1. Health service use | 45 | [19,69,106,110,116,126,127,130,143,171–206] |
| 2. Presenting complaints | 14 | [18,62,67,82,184,195,207–214] |
| 3. Coping | 21 | [22,47,84,92,105,112,114,140,176,178,206,215–224] |
| 4. Adherence | 38 | [20,22,23,27,28,33,37,47,53,58,59,70,85,86,112,116,119,133,134,140,157,167,169,188,193,197,225–236] |
| 5. Help seeking | 13 | [62,64,71,126,160,165,190,237–242] |
| 6. Initiation of treatment | 9 | [35,53,86,121,157,193,197,243,244] |
| 7. Disclosure | 9 | [42,162,231,245–250] |
| 8. Decisions concerning treatment | 1 | [88] |

We identified 105 occurrences within ‘preconception’, 128 total occurrences within ‘experience’ and 150 occurrences within ‘behavior’. The most frequently occurring single domains were ‘concept of depression’ (45), ‘health service use’ (45), ‘experience of depression’ (41), and ‘adherence’ (38). See also figure 3.

**Figure 3.**
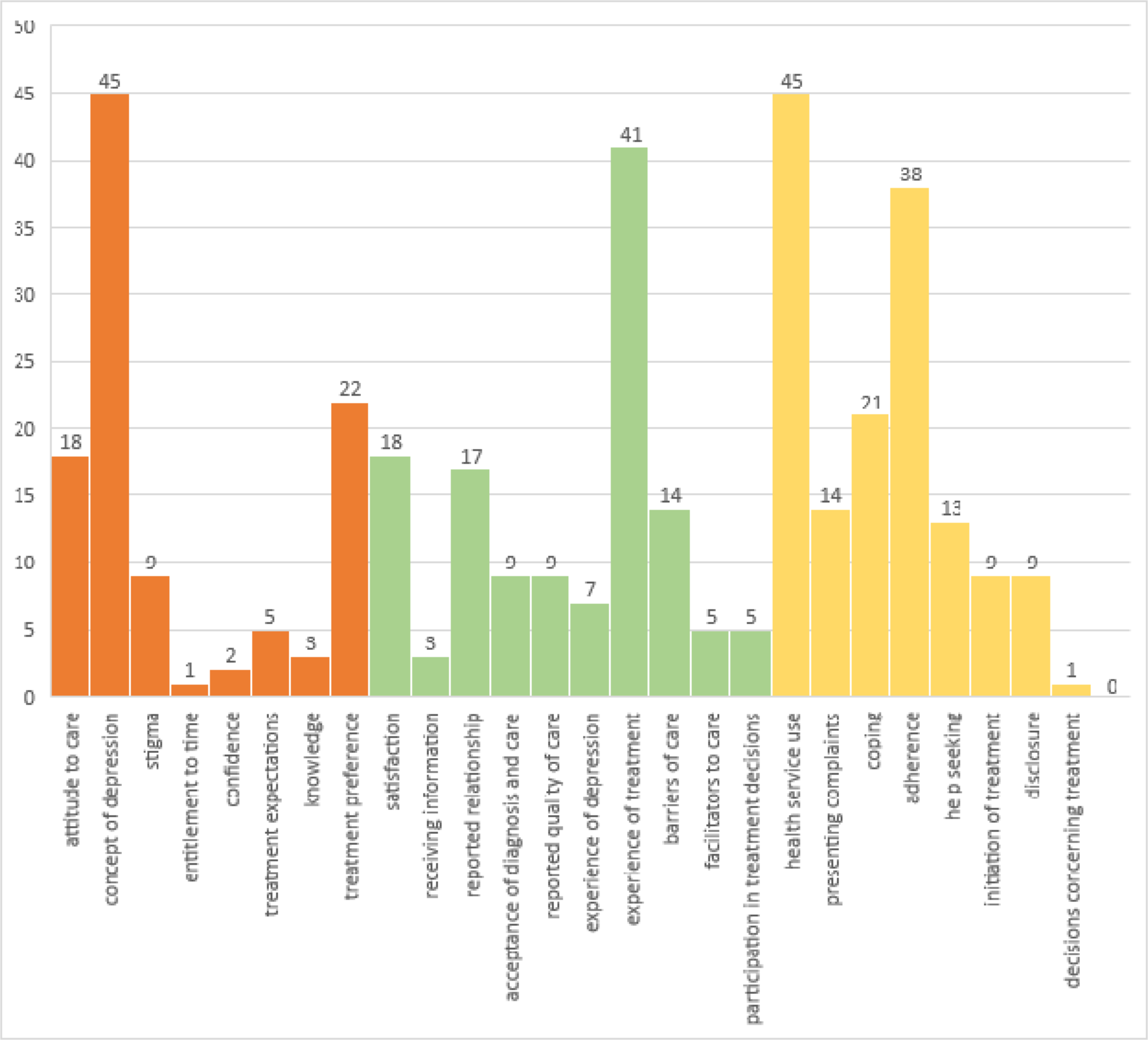
Patients’ perspectives on depression treatment in primary care: number of occurrences of domains and superordinate domains i.e., ‘preconception’ (orange), ‘experience’ (green) and ‘behavior’ (yellow).

Some terms e.g., ‘preference’, were difficult to group as a whole because of heterogenous scientific approaches. While some studies asked for treatment preferences in more naïve patients, other educated their participants thoroughly before asking to decide. Even other publications did not specify how informed their subjects were when asked for treatment preference or studied a heterogenous group in respect to that.

#### 4.5.2. Identified Barriers and Facilitators

We screened all included studies for named barriers and facilitators of treatment. We defined ‘barrier’ or ‘facilitator’ as given means by which patients’ behavior is subjectively or observably impeded or helped. Included papers necessarily included details on barriers and facilitators, which we categorized as ‘experience’, as well as behaviors they were seen in relation too. Many of the publications used for this specific synthesis also contained terms from the domain ‘preconception’ e.g., ‘stigma’ was given as a barrier to seeking help, while ‘attitude to drugs’ was commonly mentioned as impeding adherence to treatment.

Overall, we identified 61 publications testing and exploring barriers and facilitators to care. We grouped the matches using a chronological approach as to which stage of treatment is impeded o helped with i.e., diagnosis, treatment, and maintenance. Secondly, we grouped the mentioned behaviors according to our prior approach landing on ‘help-seeking’, ‘disclosure’, ‘initiation’, ‘adherence’, ‘weaning-off medication’ and ‘self-management’. Most publications (34 in total) dealt with ‘adherence’ and its barriers and facilitators. Only few publications addressed help-seeking and the maintenance of treatment. For full results see table 5.

**Table 5.** Publications on barriers and facilitators to depression treatment in primary care from the patients’ point of view.

| Treatment stage | Behavior | Description | Publication |
| --- | --- | --- | --- |
| Diagnosis | Help seeking | Time to initial medical presentation | Gormley, O'Leary, 1998 |
|  |  | Help seeking patterns | Van Hook, 1999 |
|  |  | Help seeking behaviors | Shin et al., 2002 ** |
|  |  | Help seeking | Lawrence et al., 2006 * |
|  |  | Barriers to help seeking | Callister et al., 2011 ** |
|  |  | Reasons to not present in PC with depression | Chew-Graham et al., 2012 ** |
|  |  | Help seeking pathways | Hansen et al., 2012 ** |
|  |  | Uptake of GP referral | Holloway et al., 2015 |
|  |  | Intention to seek help | Schomerus et al., 2019 |
|  | Disclosure | Informing the physician of discontinuing antidepressants | Demyttenaere et al., 2001 |
|  |  | Symptom disclosure | O'Connor et al., 2001 |
|  |  | Discuss depression | Coorigan et al., 2003 |
|  |  | Hindering disclosure | Chew-Graham et al., 2009 ** |
|  |  | Unvoiced agenda | Malpass et al., 2009 ** |
|  |  | Reasons for not disclosing depression | Bell et al., 2011 * |
|  |  | Factors hindering or facilitating disclosing symptoms | Keller et al., 2016 ** |
| Treatment | Initiation | Time to initial medical presentation | Gormley, O'Leary, 1998 |
|  |  | Perceived barriers to psychotherapy | Mohr et al., 2006-12 |
|  |  | Barriers to treatment and information | Saver et al., 2007 ** |
|  |  | Filling prescription/ | Stecker et al., 2007 |
|  |  | initiation of psychotherapy |  |
|  |  | Initiation of and adherence to treatment (escitalopram or interpersonal therapy) | Raue et al., 2009 |
|  |  | Reasons that prevent individuals from participating in a depression treatment program | Kwong et al., 2012 * |
|  |  | Factors influencing treatment uptake | Horevitz et al., 2015 * |
|  |  | Initiation of treatment | Waitzfelder et al., 2018 |
|  |  | Initiation of treatment | Cornwell et al., 2021 |
|  | Adherence | Compliance to treatment (medication) | Johnson, 1981 |
|  |  | Adherence to medication | Lin et al., 1995 |
|  |  | Adherence to treatment | Katon et al., 1995 |
|  |  | Adherence to medication | Katon et al., 2000 |
|  |  | Compliance to antidepressants | Demyttenaere et al., 2001 |
|  |  | Adherence | Brown et al., 2001 |
|  |  | Why patients stop medication | Manning, Marr, 2003 |
|  |  | Adherence to medication | Solberg et al., 2003 |
|  |  | Difficulty with continuation of care | Gask et al., 2003 ** |
|  |  | Adherence to maintenance pharmacotherapy | Lin et al., 2003 |
|  |  | Adherence to medication | Brook et al., 2005 |
|  |  | Discontinuation | Ruoff et al., 2005 |
|  |  | Adherence | Bogner et al., 2006 |
|  |  | Adherence to sertraline | Akerblad et al., 2006 |
|  |  | Medication concordance | Badger, Nolan, 2006 * |
|  |  | Adherence to medication | Brook et al., 2006 |
|  |  | Treatment adherence | Loh et al., 2007 |
|  |  | Adherence to medication | Weich et al., 2007 |
|  |  | Compliance to paroxetine | Dernovsek et al., 2008 |
|  |  | Adherence to medication | Russel, Kazantzis, 2008 |
|  |  | Filling prescriptions | van Geffen et al., 2009 |
|  |  | Compliance to antidepressants | Hérique, Kahn, 2009 |
|  |  | Completion of online-CBT | Beattie et al., 2009 * |
|  |  | Initiation of and adherence to treatment (escitalopram or interpersonal therapy) | Raue et al., 2009 |
|  |  | Initiation and continuation of antidepressant treatment | van Geffen et al., 2010 |
|  |  | Reasons for non-adherence | Fortney et al., 2011 |
|  |  | Barriers to adherence | Hansen et al., 2012 |
|  |  | Adherence | Kales et al., 2013 |
|  |  | Adherence to medication | Serrano et al., 2014 |
|  |  | Care engagement | Campbell et al., 2016 |
|  |  | Initiation of and adherence to Treatment | Vuorilehto et al., 2016 |
|  |  | Adherence to medication | Moise et al., 2017 |
|  |  | Adherence to antidepressants | Green et al., 2017 * |
|  |  | Discontinuation | Jeffery et al., 2023 |
| Maintenance | Discontinuation of medication | Barriers and facilitators to discontinuation of antidepressants | Leydon et al., 2007 ** |
|  |  | Barriers to discontinuation of antidepressants | Dickinson et al., 2010 ** |

\*: publications including qualitative data, \*\*: publications limited to qualitative data

Further we extracted, categorized, and grouped specific barriers and facilitators and synthetized them according to which behavior they are influencing. Tables 6 to 9 show the results. We assigned the extracted items to the categories ‘personal’, ‘contextual’, and ‘socioeconomic’.

**Table 6.** Barriers to help seeking. Synthesis of data drawn from publications in table 2, limited to quantitative findings.

| Domain | Barriers to help seeking | Facilitators to help seeking |
| --- | --- | --- |
| 1. Personal |  |  |
| Characteristics | N/A | N/A |
| Illness related | Neuroticism [239] | More severe depression [237] |
| Medical history | N/A | N/A |
| Attitudinal | Perceived separation between mental health and general health [237] | Desire for emotional support [238] |
|  | Stigma [237] | Stigma of psychological care [238] |
|  | Thinking that it did not affect their health [237]<br>Blaming persons with mental illness for their problem [71] | Self-identification [71]<br>More knowledge [71] |
| **2. Contextual** |  |  |
| Relationship | Not being asked [237]<br>Thinking that the staff is unhelpful and uninterested [237] | N/A |
| Treatment related | N/A | N/A |
| Interventions | N/A | N/A |
| **3. Socioeconomic** |  |  |
| Cultural background | N/A | N/A |
| Financial resources | N/A | N/A |
| Social inclusion | N/A | N/A |
| Other | Too busy [237]<br>Support for discrimination [71] | N/A |

**Table 7.** Barriers to disclosure. Synthesis of data drawn from publications in table 2, limited to quantitative findings.

| Domain | Barriers to disclosure | Facilitators to disclosure |
| --- | --- | --- |
| **1. Personal** |  |  |
| Characteristics | Female [161] | Female gender [245] |
| Illness related | More severe symptoms [161] | Higher depressive scores [161,245] |
| Medical history | No history of depression [161] | N/A |
| Attitudinal | Belief that depression is stigmatizing [161]<br>Belief that one should be in control of depression [161]<br>[Depression] not doctor's job [161]<br>[Mood] private information [161] | N/A |
| **2. Contextual** |  |  |
| Relationship | Fear of loss of emotional control [161]<br>Unsure how to introduce topic [161]<br>Fear to distract doctor [161]<br>Fear of being thought less of [161] | Better relationship [231] |
| Characteristics of PCP | N/A | N/A |
| Treatment related | Medication aversion [161]<br>Medical records possibly unsafe [161]<br>Fear of psychiatric or counselling referral [161]<br>Fear of being a 'psychiatric patient' | Previous contact with a psychiatrist [245] |
|  | [161] |  |
| Interventions | N/A | N/A |
| **3. Socioeconomic** |  |  |
| Cultural background | Hispanic [161] | N/A |
| Financial resources | Less income [161] | N/A |
| Social inclusion | Satisfaction with others [247]<br>Better social support [247] | N/A |
| Other | Less education [161] | N/A |

**Table 8.** Barriers to initiation. Synthesis of data drawn from publications in table 2, limited to quantitative findings.

| Domain | Barriers to initiation | Facilitators to initiation | Investigated without significant results |
| --- | --- | --- | --- |
| **1. Personal** |  |  |  |
| Demographics | Asian, African-American or Hispanic [243]<br>Senior patients [243,244]<br>Male [244] | Male [243]<br>Hispanic [244] | N/A |
| Illness related | Lower baseline PHQ-2 [244] | More severe depression [243] | N/A |
| Medical history | N/A | Fewer comorbidities [243] | N/A |
| Attitudinal | Discomfort talking about personal issues with someone unknown [159]<br>Concerns what others might think [159] | Strong preference for received treatment [86] | Stigma [35]<br>Belief that psychotherapy weakens the chance of obtaining a job [35] |
| Other | More neuroticism [239]<br>Prior mental health diagnosis [244] | Prior mental health treatment [243] | N/A |
| **2. Contextual** |  |  |  |
| Relationship | 'Warm handoff' (for Spanish-speaking Latinos) [121] | N/A | N/A |
| Characteristics of PCP | N/A | N/A | N/A |
| Treatment related | Cost of psychotherapy [159]<br>Prior visit to clinic [244] | Same day mental health appointment [244] | N/A |
| Interventions | N/A | N/A | N/A |
| **3. Socioeconomic** |  |  |  |
| Cultural background | N/A | N/A | N/A |
| Financial resources | N/A | N/A | N/A |
| Social inclusion | Married [244] | N/A | N/A |
| Other | Time constraints [159] | N/A | N/A |
|  | Transportation difficulties [159]<br>Care for child or other [159] |  |  |

**Table 9.**
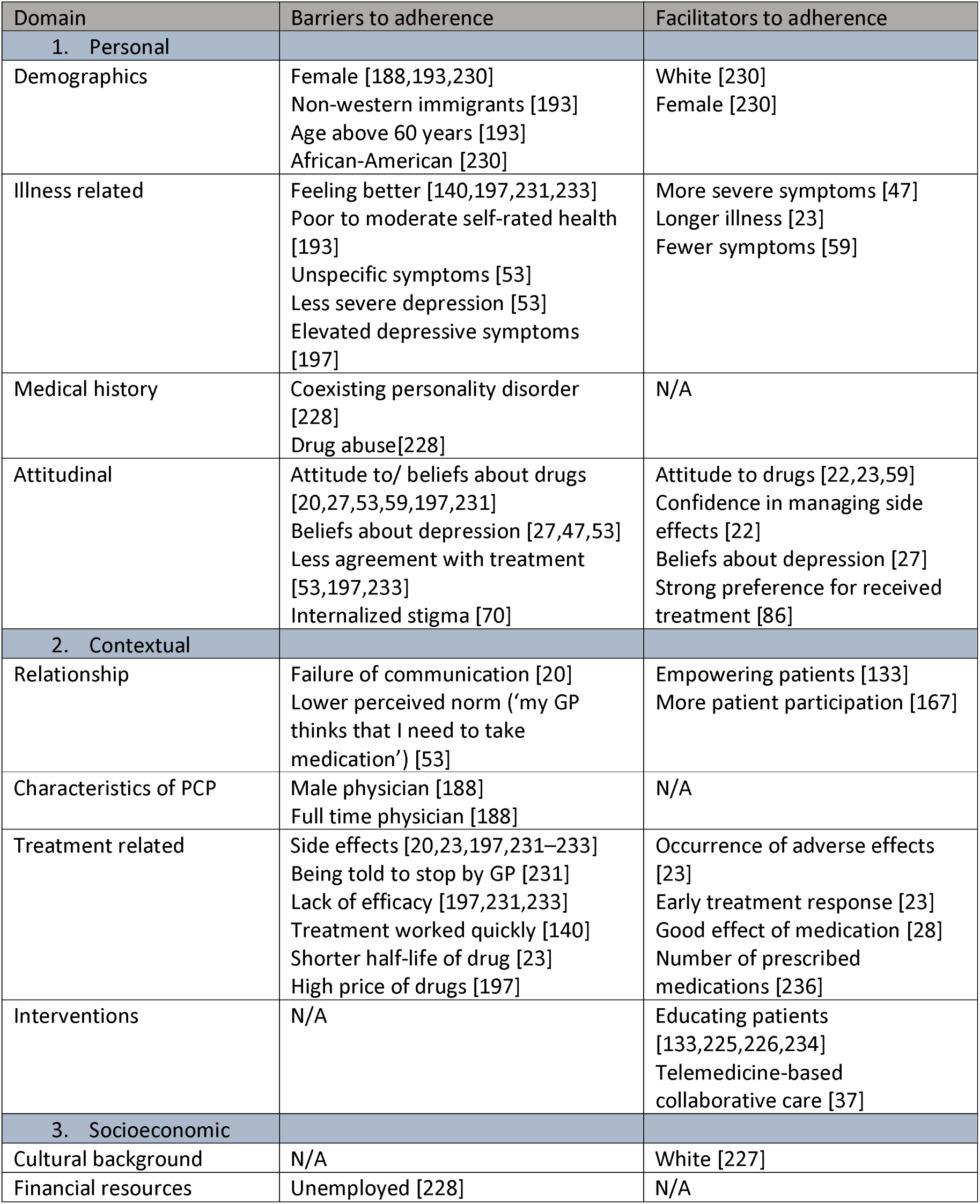
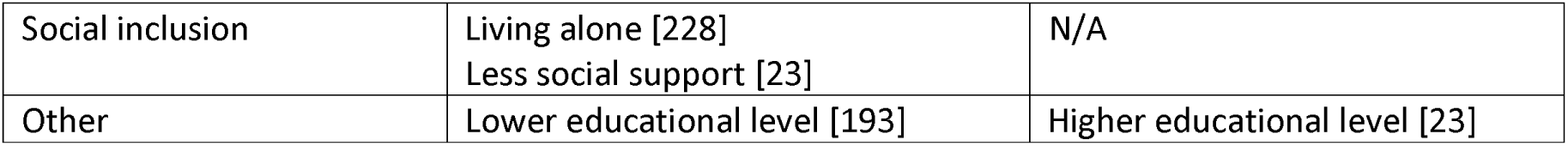
Barriers to adherence. Synthesis of data drawn from publications in table 2, limited to quantitative findings.

Table 5 contains all publications dealing with barriers and facilitators. For the synthesis in table 6 to 9 we only included quantitative results from mentioned publications. To comprehensively extract qualitative findings conducting a detailed qualitative meta-synthesis would have been needed.

Overall, most barriers and facilitators were grouped as either illness- or treatment-related or attitudinal. Socioeconomic factors, the impact of practitioners’ characteristics and the doctor-patient relationship were less researched.

#### 4.5.3. Presenting Complaints

Building on the concept of ‘barriers and facilitators’ we looked for other relevant topics that might influence treatment. We decided to take a closer look at presenting complaints as to which symptoms patients report when depression is present or even diagnosed. While psychological complaints such as ‘low mood’ point practitioners into the right direction, the primary presentation of physical symptoms might on the other hand impede diagnosis.

We included all publications on ‘presenting complaints’ with the exception of two [208,210] which did not report the relevant data in a manner that appeared useful for the synthesis. Tylee et al. studied number and timing symptom report within the consultation [214]. We included the publication into the following table for an overview over the methodology without extracting the ‘percentage of somatic complaints’.

Due to our inclusion criteria, all outcomes for this category derived from open-ended questions. The reported symptoms were further categorized by the authors. We defined ‘somatic complaints’ as physical, vegetative, or somatic symptoms. Table 10 provides an overview of included publications and extracted data.

**Table 10.** Publications covering presenting complaints.

| Publication | Outcome | Number of Participants | Results | Percentage of somatic complaints |
| --- | --- | --- | --- | --- |
| Widmer et | Chief | 154 | At time of diagnosis 23% of depressed | 64 |
| al., 1978 | complaint |  | patients gave low mood, 33% sleep disturbances, 31% fatigue, 4% anxiousness or agitation and 9% other symptoms as their chief complaint. |  |
| Diamond et al., 1987 | Reason for visit | 67 | The reasons for visit given by patients were 34,5% somatic complaints, 19,5% vegetative and 21% psychological. | 60 |
| Williamson et al., 1989 | Initial complaint | 7 | In the family practice center depressed patients gave following initial complaints: "General yuck", Physical exam, Kidney problem, Injured finger, Severe headaches, "Multiple complaints", Arthritis. | 100 |
| Kirmayer et al., 1993 | Presenting complaints | 202 | Of the patients with CES-D scores of 16 or higher, 14% (N=31) were psychosocial presenters, 32% (N=69) were initial somatizers, 24% (N=52) were facultative somatizers, and 23% (N=50) were true somatizers. The presentations of 6% (N=13) were classified as health maintenance visits. These patients were excluded from further comparisons. Facultative and true somatizers only reported physical complaints. | 85*/50 |
| Tylee et al., 1995 | Mentioning of symptoms | 72 (women) | A total of 13 mentioned between 10 and 19 physical symptoms and four patients mentioned between 20 and 29 physical symptoms. The first mention of a psychiatric symptom was within the first four mentions of any symptoms for 30 women. 41 women first mentioned a psychiatric symptom after the first four mentions or mentioned no psychiatric symptom. | N/A |
| Cornwell et al., 1998 | Presenting complaints | 90 (40 South Asian, 50 White persons) | Within the South Asian group 27 (67,5%) presented with physical complaints, 38 (95%) presented with psychological complaints (n=40). In the White group 11 (22%) presented with physical complaints, 50 (100%) presented with psychological complaints (n=50). | 68*/ 22* |
| Waza et al., 1999 | Symptom report | 189 (85 USA, 104 Japan) | USA (N=85): 74.1% also reported physical symptoms, 9.4% reported only physical symptoms, 16.5% reported only psychological symptoms. Japan(n=104): 69.3% also reported physical symptoms, 26.9% reported only physical symptoms, 3.8% reported only psychological symptoms. | 74*/ 69* |
| Mckelvey et | Presenting | 1057 | Of the patients with a CES-D above the | 77 |
| al., 2001 | complaints |  | cut-off 77% (813) presented with physical complaints, 23% (244) presented with psychological complaints. |  |
| Yeung et al., 2004 | Presenting complaints | 29 | The most common presenting complaints were fatigue (N=5), insomnia (N=5), headache (N=4), cough (N=2), pain (N=2), dizziness (N=2), cervical problems (N=1), and sexual dysfunction (N=1). Four complained of psychological symptoms, 2 described feelings of nervousness. One did not spontaneously report any symptoms. | 76 |
| Menchetti et al., 2009 | Reason for consultation | 250 | 110 depressed patients (44.0%) consulted their PCP for psychological or family problems, 102 subjects (40.8%) for physical illness, and 38 subjects (15.2%) for pain symptoms. | 56 |
| Chen et al., 2015 | Chief complaint | 190 | Subjects were most likely to report a chief complaint in the category of Depressed mood/ unhappiness/mood problems (52.1%), followed by Psychosocial stressors (38.9%), Depressive neurovegetative symptoms (31.6% — e.g., poor sleep, poor appetite, low energy), and Depressive psychological symptoms (24.2% — e.g., rumination, difficulty concentrating, agitation). Multiple responses were allowed. | 32* |
| Heinz et al., 2021 | Reporting complaints | 430 | 190 (44.2%) patients with depression reported physical complaints only, 42 (9.8%) reported mental complaints only, 124 (28.8%) consulted the GP due to mental and physical complaints, 19 (4.4%) stated no complaints and 55 (12.8%) gave other reasons. | 44/ 29* |

\* Percentage of participants also, not necessarily exclusively, reporting physical complaints.

Between 44 and 100% of studied patients presented only with physical symptoms. The percentage of patients also reporting physical symptoms when seeking help for depression was given between 22 and 85%. Some publications also studied the correlation between depression state and number of symptoms given, which we did not further synthesize [82,214].

## 5. Discussion

### 5.1. Characteristics of publications and overall findings

Even though depression is very common and often treated exclusively in the primary care context, still most research towards depression care is conducted in specialized contexts [251]. Looking at the patients’ perspective of care beyond symptoms is sometimes a secondary topic, when conducting a randomized-controlled-trial, but has also been a researched outcome since at least the 1970s. The majority of publications related to our research questions were published in the early 2000s. Including patients as important stakeholders not just as subjects but also in planning and conducting research has been a growing trend in recent years [252]. It is to be expected that this will lead to more publications on the patient’s perspective in years to come.

Even though we limited our findings to OECD member states for the screening process it is still notable that most publications stemmed from the Anglo-American region. Our research zooms in on a very distinct context that depends on not only individual but also social on political parameters. We cannot say if findings from the US apply for German primary care settings or vice versa. On the other hand, primary care itself and its research is a heterogenous field with a multitude of settings and scientific approaches.

Based on the literature we provide a comprehensive framework for the concept ‘patients’ perspective’ which can be used for future studies, applied internationally, and be built on creating networks and foci for research and intervention. The implementation of the patients’ perspectives in future guidelines and policy is essential to work towards more accessible care for all patients with depression. The primary care context provides an ideal setting for impactful research and intervention [253].

### 5.2. Patients’ perspectives on depression treatment in primary care

With this work we provide an overview of current literature on patients’ perspectives on depression treatment in primary care. We defined and evaluated a structured approach to the concept trying to grasp its width and depth. By evaluating the method throughout the entire process, we ensured an objective and understandable approach of literature synthesis.

We identified and grouped domains we consider relevant to the concept of patients’ perspective. The premeditated superordinate domains seemed useful for that process.

Comparing superordinate domains, their terms, and their numbers of occurrences, we found that research approaches and scientific research towards them varies. Behavioral terms have been studied since at least the 1970s and provide clear comparable quantitative measures. The fact that patients’ behavior is observable and close to a physician-centered approach is a possible explanation.

Apart from behavioral terms we found that ‘satisfaction’ and ‘preference’ are commonly studied outcomes. They rarely constitute main outcomes of studies but are easily retrievable data derived from longitudinal and cross-sectional studies. After quality appraisal these publications can be included in separate systematic reviews. Based on our review, we recommend that these outcomes should be included in intervention studies. We also recommend to precisely state how these outcomes were generated to provide accessible and comparable data for evidence synthesis and future research.

Surprisingly, we did not find many publications researching patients’ expectations [44,86]. Even though many terms grouped in ‘preconception’ are related to ‘expectation’ it is rarely directly addressed in studies. Terms like ‘acceptability’, ‘preference’ and ‘satisfaction’ are possibly dependent on expectations and could be predicted by them. Further research could close that gap and lead to an early intervention instead of hindsight, as measured by ‘satisfaction’. Current research stresses the link between patient-provider-relationship and satisfaction [254].

The terms ‘experience of depression’ and ‘experience of treatment’ are broad and more complex to be synthesized. Publications on those terms used a qualitative approach. We did not perform a qualitative meta synthesis, which would be an interesting next step regarding these domains.

### 5.3. Barriers and facilitators to treatment faced by patients

According to our definition of ‘barriers and facilitators’ i.e., conditions and interventions impeding and helping with care, we identified and grouped related topics. ‘Barriers and facilitators’ showed an overlap between all three superordinate domains. While by our definition closely linked to behavior, we categorized ‘barriers and facilitators’ themselves as ‘experience’. Certain terms we categorized as ‘preconception’ were also identified as specific barriers e.g., ‘stigma’ and ‘attitude to care’.

For scoping, we found this framework to be adequately clear and inclusive.

Most research focused on adherence and lack thereof, often contributing as a secondary outcome to large-scaled medication studies. While drug-associated barriers like ‘side effects’ may not be specific to the primary care setting, barriers and facilitators describing the patient-provider-relationship may be. This has been shown for other medical issues and drugs [255].

Preference for the prescribed treatment [86], participation [167], and education [234] have been described as helpful to improve patients’ motivation for and therefore adherence to care. Nevertheless, more research is needed to build from these findings.

We did not find research on geographic and other community factors as a barrier to help-seeking and care, even though we believe it to be a challenge faced especially by under-served communities, who might be underrepresented in current research [256]. Among our inclusions, socioeconomic factors are less researched than personal and contextual factors.

Overall, factors that were not directly related to treatment and illness e.g., attitudinal or relationship aspects, were less researched. With the rise of integrated concepts like collaborative care [257], we believe these aspects as important to be included into future research and policy.

Among our included publications there are also other topics arising that are related to barriers and facilitators to care even though they do not clearly frame it that way. Not just if patients disclose symptoms but also the type of presenting complaints influences if they are diagnosed correctly.

Among publications studying the quality and quantity of complaints there is a trend towards patients reporting physical symptoms. Three of the identified studies correlated the quality (physical vs. psychological), quantity and timing of complaints with the correct recognition of the depressive state by the GP and found that the report of physical symptoms as well as the delayed report of psychological symptoms reduce the chances of being diagnosed with depression [195,213,214]. Reporting mainly physical complaints might also be a contextual effect. Williamson et al. compared patients visiting a family practice with those consulting a psychiatric clinic and found this to be a distinction i.e., primary care patients reporting more somatic symptoms [209]. Other contextual effects such as sociocultural aspects, national and regional, might also play a role on patients’ symptom report [18]. It is important to further research presenting complaints and behaviors with the questions whether and to what degree they can impede proper diagnosis.

Data about patient-sided barriers and facilitators towards depression treatment in primary care still seems inconclusive with contradictory and heterogenous approaches and results. More research and synthesis are needed for which we provided a framework.

### 5.4. Strengths and limitations

To our knowledge, this is the first literature review on patients’ perspectives on depression treatment in primary care offering a comprehensive framework for future studies.

We conducted a premeditated and thorough search for all relevant studies. Because the findings were limited to studies set in the primary care context, we naturally excluded other designs and studies that may have contributed to the question of patients’ perspectives, especially preconceptions.

We limited our search to MEDLINE and Psycinfo, not specifically looking for grey literature. We believe our data to be representative of current research and our framework to be applicable for possibly missed publications.

We still think that the identified research topics point out important factors for treatment. We conceptualized the abstract term ‘perspective’ for the purpose of our research and found it sufficiently broad and specific. We do not claim our concept of ‘perspective’ to be the only valid one. We encourage future investigators to critique and build from our concept.

In the planning stage of this project, we were advised on its design by patients from the *Munich Alliance Against Depression* (Münchner Bündnis gegen Depression e.V.) an interest group organized in Germany for patients with a history of depression.

## 6. Conclusion

This scoping review conceptualizes ‘patients’ perspective on depression treatment in primary care’ as a scientific field with its domains, subdomains, and research approaches and provides a comprehensive overview of current literature, providing a framework for future studies. Patients’ behavioral measures are an easy way to implement patients’ perspectives as a secondary outcome into prospective and cross-sectional study designs.

Findings about specific barriers and facilitators of care which are already described and studied are summarized as well as grouped in an approachable and systematic manner to help guide future research. While illness- and treatment-related barriers and facilitators are commonly researched, there is less data on how the patient-provider-relationship impacts patient behavior. While adherence is focused on most commonly, there is little evidence on what hinders or encourages help-seeking, disclosure, or maintenance. Patients often present with physical symptoms when seeking help for depression which can hinder or delay diagnosis.

Patients’ perspectives on depression treatment in primary care are an essential aspect of the maintenance and improvement of evidence-based care and should be included in guidelines and policies.

## 7. Clinical implications

Even though there is a trend towards more patient participation in research [258], clinical practice guidelines still rarely consult patients and their experiences in the process of making decisions on recommendations and implementation.

The purpose of this framework is three-fold: First, it can be used as a base for questionnaires researching the concept of patients’ perspectives on depression treatment in primary care. Quantifiable measures, such as satisfaction, adherence, or preference, are easy to integrate into clinical trials and offer the possibility for comparison and meta-analysis.

Second, patient-related measures about their experience and expectations can help to guide policies, to validate and evaluate models of quality of care, to improve guideline development processes, and consequently can make treatment more patient-centered, targeted and efficient. Effective implementation of guidelines and changes in heuristics in primary care require research that includes perspectives of all relevant stakeholders along with contextual factors. To create and test tailored implementation strategies it is crucial to comprehensively understand contextual barriers and facilitators [259]. Especially barriers to help-seeking are an important gap. A better understanding of help-seeking and facilitating interventions seems promising to help with early diagnosis in primary care, prevent chronicity, and reduce the burden of depression.

Third, the synthesis of barriers and facilitators offers a theoretical base on which networks can be built to better understand what the possible targets for intervention are. In particular, an understanding of contextual barriers from the patients’ point of view can help to tailor effective implementation strategies.

Figure 4 shows a theoretical model of how patients’ perspectives on care can be included into the forming and implementation of clinical practice guidelines to help with evidence-based and effective treatment of depression in primary care.

**Figure 4.**
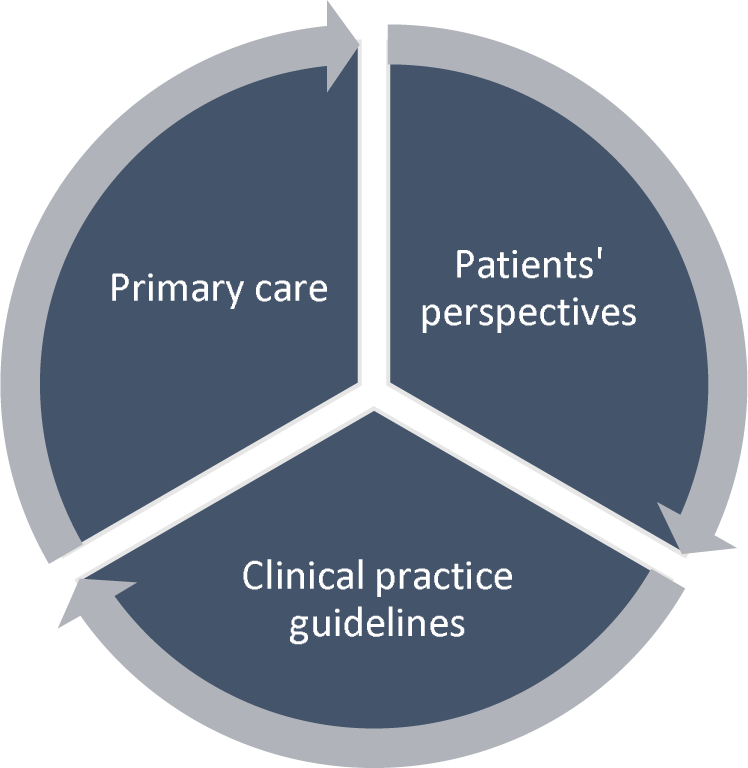
Model of impact of patients’ perspectives on clinical practice guidelines, primary care, and vice-versa.

## Data Availability

All relevant data for the evidence synthesis is given in the appendix. Complete search and review files (Rayyan export files) can be acquired by emailing the corresponding author.

https://osf.io/p9rnc

## Acknowledgements

The POKAL-Group (PrädiktOren und Klinische Ergebnisse bei depressiven ErkrAnkungen in der hausärztLichen Versorgung (POKAL, DFG-GrK 2621)) consists of the following principle investigators: Tobias Dreischulte, Peter Falkai, Jochen Gensichen, Peter Henningsen, Markus Bühner, Caroline Jung-Sievers, Helmut Krcmar, Karoline Lukaschek, Gabriele Pitschel-Walz and Antonius Schneider. The following doctoral students are members of the POKAL-Group: Katharina Biersack, Constantin Brand, Vita Brisnik, Christopher Ebert, Julia Eder, Feyza Gökce, Carolin Haas, Lisa Hattenkofer, Lukas Kaupe, Jonas Raub, Philipp Reindl-Spanner, Hannah Schillok, Petra Schönweger, Clara Teusen, Marie Vogel, Victoria von Schrottenberg, Jochen Vukas and Puya Younesi.

S 1 Appendix

S2 Preferred Reporting Items for Systematic reviews and Meta-Analyses extension for Scoping Reviews (PRISMA-ScR) Checklist

