## Supplementary material for "Beyond the medical file: a scoping review on patients’ perspectives on guideline-oriented depression treatment in primary care": S1 Appendix

Appendix 1: Search strategy

| Medline Search, conducted 08/02/2023 |
| --- |
| ("Attitude"[MeSH Terms] OR "Attitude to Health"[MeSH Terms] OR ("knowledge"[MeSH Terms] OR "knowledge"[All Fields] OR "knowledge s"[All Fields] OR "knowledgeability"[All Fields] OR "knowledgeable"[All Fields] OR "knowledgeably"[All Fields] OR "knowledges"[All Fields]) OR ("Help"[All Fields] AND ("seeking"[All Fields] OR "seeks"[All Fields])) OR "Self-Management"[MeSH Terms] OR "adaptation, psychological"[MeSH Terms] OR ("illness behaviour"[All Fields] OR "illness behavior"[MeSH Terms] OR ("illness"[All Fields] AND "behavior"[All Fields]) OR "illness behavior"[All Fields]) OR ("experience"[All Fields] OR "experience s"[All Fields] OR "experiences"[All Fields]) OR ("qualities"[All Fields] OR "quality"[All Fields] OR "quality s"[All Fields]) OR "Role"[MeSH Terms] OR "Self Concept"[MeSH Terms] OR "Self Efficacy"[MeSH Terms] OR ("belief s"[All Fields] OR "culture"[MeSH Terms] OR "culture"[All Fields] OR "belief"[All Fields] OR "beliefs"[All Fields]) OR ("believability"[All Fields] OR "believable"[All Fields] OR "believed"[All Fields] OR "believer"[All Fields] OR "believers"[All Fields] OR "believing"[All Fields] OR "culture"[MeSH Terms] OR "culture"[All Fields] OR "believe"[All Fields] OR "believes"[All Fields]) OR (("illness"[All Fields] OR "illness s"[All Fields] OR "illnesses"[All Fields]) AND ("percept"[All Fields] OR "perceptibility"[All Fields] OR "perceptible"[All Fields] OR "perception"[MeSH Terms] OR "perception"[All Fields] OR "perceptions"[All Fields] OR "perceptional"[All Fields] OR "perceptive"[All Fields] OR "perceptiveness"[All Fields] OR "percepts"[All Fields])) OR (("patient s"[All Fields] OR "patients"[MeSH Terms] OR "patients"[All Fields] OR "patient"[All Fields] OR "patients s"[All Fields]) AND ("perspective"[All Fields] OR "perspective s"[All Fields] OR "perspectives"[All Fields])) OR ("coherence"[All Fields] OR "coherences"[All Fields] OR "coherencies"[All Fields] OR "coherency"[All Fields] OR "coherent"[All Fields] OR "coherently"[All Fields]) OR (("therapeutics"[MeSH Terms] OR "therapeutics"[All Fields] OR "treatments"[All Fields] OR "therapy"[MeSH Subheading] OR "therapy"[All Fields] OR "treatment"[All Fields] OR "treatment s"[All Fields]) AND "inititation"[All Fields]) OR ("expect"[All Fields] OR "expectable"[All Fields] OR "expectance"[All Fields] OR "expectant"[All Fields] OR "expectative"[All Fields] OR "expected"[All Fields] OR "expecting"[All Fields] OR "expects"[All Fields] OR "motivation"[MeSH Terms] OR "motivation"[All Fields] OR "expectancies"[All Fields] OR "expectancy"[All Fields] OR "expectation"[All Fields] OR "expectations"[All Fields]) OR ("expect"[All Fields] OR "expectable"[All Fields] OR "expectance"[All Fields] OR "expectant"[All Fields] OR "expectative"[All Fields] OR "expected"[All Fields] OR "expecting"[All Fields] OR "expects"[All Fields] OR "motivation"[MeSH Terms] OR "motivation"[All Fields] OR "expectancies"[All Fields] OR "expectancy"[All Fields] OR "expectation"[All Fields] OR "expectations"[All Fields]) OR ("expect"[All Fields] OR "expectable"[All Fields] OR "expectance"[All Fields] OR "expectant"[All Fields] OR "expectative"[All Fields] OR "expected"[All Fields] OR "expecting"[All Fields] OR "expects"[All Fields] OR "motivation"[MeSH Terms] OR "motivation"[All Fields] OR "expectancies"[All Fields] OR "expectancy"[All Fields] OR "expectation"[All Fields] OR "expectations"[All Fields]) OR "Patient Preference"[MeSH Terms] OR ("decision making, shared"[MeSH Terms] OR ("decision"[All Fields] AND "making"[All Fields] AND "shared"[All Fields]) OR "shared decision making"[All Fields] OR ("shared"[All Fields] AND "decision"[All Fields] AND "making"[All Fields])) OR ("physical examination"[MeSH Terms] OR ("physical"[All Fields] AND "examination"[All Fields]) OR "physical examination"[All Fields] OR "physical"[All Fields] OR "physically"[All Fields] OR "physicals"[All Fields]) OR "somat*"[All Fields] OR "non-specific"[All Fields] OR ("complaint"[All Fields] OR "complainted"[All Fields] OR "complaints"[All Fields]) OR ("psychosomatic"[All Fields] OR "psychosomatical"[All Fields] OR "psychosomatically"[All Fields] OR "psychosomatics"[All Fields] OR ("psychophysiologic disorders"[MeSH Terms] OR ("psychophysiologic"[All Fields] AND "disorders"[All Fields]) OR "psychophysiologic disorders"[All Fields])) OR ("pain"[MeSH Terms] OR "pain"[All Fields]) OR ("diagnosis"[MeSH Subheading] OR "diagnosis"[All Fields] OR "symptoms"[All Fields] OR "diagnosis"[MeSH Terms] OR "symptom"[All Fields] OR "symptom s"[All Fields] OR |

| |
|---|
| "symptomes"[All Fields])) AND ("General Practice"[MeSH Terms] OR "General Practitioners"[MeSH Terms] OR "Family Practice"[MeSH Terms] OR "Primary Health Care"[MeSH Terms] OR "physicians, primary care"[MeSH Terms] OR "Ambulatory Care"[MeSH Terms]) AND ("Depressive Disorder"[MeSH Terms] OR "depressive disorder, major"[MeSH Terms] OR "Depression"[MeSH Terms]) |
| Psycinfo Search, conducted 08/02/2023 |
| (general practice or GP or family practice or primary health care or physicians primary care or ambulatory care)<br>AND<br>(depression or depressive disorder or depressive symptoms or major depressive disorder)<br>AND<br><br>(attitude or knowledge or help seeking or self-management or adaptation or illness behavior or experience or belief or illness perception or patient perspective or coherence or treatment inititation or expect or shared decision making or patient preference or symptom or complaint) |

2

3 Appendix 2:

| Authors/ Year | Study design | Number of partici-pants | Population: Definition of depression | Population: Special Group | Context: Region | Context: Primary care | Concept |
|---|---|---|---|---|---|---|---|
| Johnson, 1973 | Longitudinal, Interview | 73 | Diagnosis by GP | . | UK | GP | Number of consultations<br>Satisfaction<br>Relationship |
| Widmer, 1978 | Longitudinal, Case-control-matching, Patient records | 154 | Diagnosis by GP | . | US | FP | Consultation frequency<br>Chief complaint |
| Widmer RB, Cadoret RJ, 1979 | Case-control-matching, Patient records | 43 | Diagnosis by GP | . | US | FP | Consultation frequency |
| Cadoret et al., 1980 | Case-control-matching, Medical records | 234 | Diagnosis by GP | . | US | FPC | Consultation frequency |
| Johnson DA, 1981 | Longitudinal, Interview | 117 | BDI ≥ 11 | . | UK | GP | Compliance<br>Attitude to medication<br>Doctor-patient-communication |
| Cremniter et al., 1982 | Cross-sectional, Survey | 682 | Diagnosis by GP | . | F | GP | Presenting symptoms |
| Wilson et al., 1983 | Longitudinal, Case-control-matching, Record audit | 101 | Diagnosis by GP | . | US | FPC | Consultation frequency |
| Oxman et al., 1983 | Cross-sectional | 126 | Chart diagnosis | . | US | FPC | Number of calls |
| Katon et al., 1983 | Cross-sectional | 147 | Zung depression test | . | US | Family medical center | Medical utilization |
| Foster JM, Gallagher D, 1986 | Case-control-matching, Health and Daily Living Questionnaire | 32 | Diagnosis by GP | Elderly | US | VC | Coping |
| Diamond et al., 1987 | Longitudinal, Patient charts | 67 | Diagnosis by GP | Women | US | FP | Reason for visit |

| | | | | | | | |
|---|---|---|---|---|---|---|---|
| Williamson PS, Yates WR, 1989 | Cross-sectional, Review of symptoms, Patient chart | 105 | Inventory to diagnose depression, DSM III | . | US | FPC | Presenting complaints |
| Romans-Clarkson et al., 1990 | Cross-sectional, Interview | 314 | Present State Examination (PSE) | Women | NZ | GP | Number of visits to GP Help seeking |
| Katon et al., 1990 | RCT, Physician Review Form | 124 | National Institute of Mental Health Diagnostic Interview Schedule, DSM-III-R | . | US | PCC | Health care utilization |
| Gerber et al., 1992 | Cross-sectional, Physician Recording Form | 1055 | HSCL-90 Depression Scale≥7 | . | US | GP | Presenting complaints |
| Scott AI, Freeman CP, 1992 | RCT, Questionnaire | 101 | DSM-III, diagnostic interview | . | UK | GP | Satisfaction |
| Kirmayer et al., 1993 | Observational, Interview | 202 | DIS, CES-D ≥16 | . | Canada | FMC | Clinical presentation |
| Blanchard et al., 1994 | Follow-up, Interview | 96 | Short-CARE, GMS-AGECAT | Older people | UK | GP | Declaration of symptoms |
| Callahan et al., 1994 | Longitudinal, Interview | 1711 | CES-D ≥16 | Elderly | US | GP | Health service use |
| Karlsson et al., 1995 | Case control study | 562 | Present state examination interview | . | Finland | Primary care station | Frequent attendance |
| Lin et al., 1995 | Longitudinal, Interview | 155 | Antidepressant prescription for depression | . | US | PCP | Adherence |
| Katon et al., 1995 | RCT, Questionnaire | 193 | Diagnosis by GP | . | US | PCP | Adherence Quality rating |
| Tylee et al., 1995 | Observational, Interview, Analyzed video-recordings | 72 | Diagnostic interview | Women | UK | GP | Mentions of symptoms |

| | | | | | | | |
|---|---|---|---|---|---|---|---|
| Brown et al., 1996 | RCT,<br>Health Locus of Control Scale | 272 | CES-D ≥ 22, DIS | African American | US | PCP | Health locus of control |
| Gormley N, O'Leary D, 1998 | Cross-sectional,<br>Interview | 100 | HRSD ≥16 | . | Ireland | GP | Time to presentation |
| Cornwell J, Hull S, 1998 | Cross-sectional,<br>Data from records | 90 | Antidepressant prescription for depression | South Asian | UK | GP | Presenting complaints |
| Rubio Montañés et al., 1998 | Case-control-matching,<br>Questionnaire | 209 | Escala de Ansiedad y Depresión de Goldberg (EADG) | . | Spain | HCC | Over-attendance |
| Van Hook, 1999 | Cross-sectional,<br>Questionnaire | 321 | Medical Outcome Services (MOS), ICD-10 | Women | US | PCC | Help seeking patterns |
| Kessler et al., 1999 | Cross-sectional,<br>Questionnaire | 305 | General Health Questionnaire ≥3 | . | UK | GP | Symptom interpretation |
| Arve et al, 1999 | Cross-sectional | 847 | Zung depression test | 70 years and older | Finland | GP | Self-perception of depression |
| Waza et al., 1999 | Observational,<br>Chart audit | 189 | Chart diagnosis | . | Japan/ US | FP | Complaints |
| Gilmore KA; Hargie O; 2000 | Cross-sectional,<br>Interview | 8 | Prior diagnosis | . | UK | GP | Satisfaction |
| Rost et al., 2000 | Cross-sectional,<br>Interview | 240 | Inventory to diagnose depression | . | US | PCP | Acceptability |
| Nichols GA, Brown JB, 2000 | Cross-sectional,<br>Questionnaire | 1161 | Positive screen/ chart diagnosis | . | US | FP | Being asked |
| Cooper et al., 2000 | Cross-sectional,<br>Survey | 76 | CESD ≥16 | . | US | PCC | Attitude to care<br>Importance of different aspects of care |
| Katon et al., 2000 | RCT,<br>Automated databases | 1599 | SCID | . | US | PCC | Consultation frequency<br>Adherence |

| | | | | | | | |
|---|---|---|---|---|---|---|---|
| Dwight-Johnson et al., 2000 | Cross-sectional, Interview | 1187 | CIDI | . | US | PCC | Preference |
| Bedi et al., 2000 | Partially randomised preference trial, Preference between antidepressants or psychotherapy given by patient, Questionnaire | 323 | Diagnosis by GP | . | UK | GP | Preference<br>Attitude to treatment |
| Demyttenaere et al., 2001 | Longitudinal, Antidepressant Compliance Questionnaire | 272 | Diagnosis by GP | . | US | GP | Compliance<br>Informing physician |
| Andrew et al., 2001 | Case-control-matching, Primary care records | 90 | Seasonal affective Disorder (SPAQ), DSM IV | . | UK | GP | Consultation rate |
| Meredith et al., 2001 | Cross-sectional, Components of Primary Care Index, Medical Outcomes Study, Consumer Assessment of Health Plans Survey, Data from patient surveys | 1104 | CIDI | . | US | primary care visitors in managed care organizations | Satisfaction<br>Relationship<br>Quality of care indicators |
| Rogers et al., 2001 | Qualitative interviews | 27 | Diagnosis by GP | . | UK | GP | Experience of care |
| Brown et al., 2001 | Cross-sectional, Brief COPE Inventory, Consumer Assessment of Health Plans Survey, Modified Illness Perceptions Questionnaire | 41 | Diagnosis by GP | . | US | PCC | Coping<br>Adherence<br>Illness model<br>Controllability |

| | | | | | | | |
|---|---|---|---|---|---|---|---|
| Andrews G, Carter GL, 2001 | Cross-sectional, Interview, UK Survey of Psychiatric Morbidity | 10641 | CIDI | . | Australia | GP | Number of consultations<br>Perceived need for care<br>Report of received treatment |
| Dwight-Johnson et al., 2001 | RCT, Patient assessment questionnaire | 1187 | CIDI | . | US | PCC | Preference |
| O'Connor et al., 2001 | Cross-sectional, Interview | 1021 | Diagnosis by GP | Elderly patients | Australia | GP | Symptom disclosure |
| McKelvey et al., 2001 | Cross-sectional, Summary sheets | 2792 | CES-D, DSI-SS | . | US | GP | Chief complaints |
| Webster et al., 2001 | Cross-sectional, Questionnaire | 574 | EPDS | Women postpartum | Australia | GP | Visiting health care provider<br>Satisfaction |
| Ronalds et al., 2002 | Follow-up, Interview | 148 | Psychiatric Assessment Schedule | . | UK | GP | Consultation rate |
| Holopainen D, 2002 | Cross-sectional, Interview | 7 | Recruited from support group | Women postpartum | Australia | GP | Experience of seeking help |
| Shin et al., 2002 | Focus group | 70 | Self-identification as depressed | Korean immigrants | US | PC | Help seeking |
| Maidment et al., 2002 | Cross-sectional | 67 | Antidepressant prescription for depression | 65 and older | UK | GP | Adherence (Global adherence measure (GAQ))<br>Patient education (the questionnaire on patient education (QPE))<br>Insight (schedule for assessing the three components of insight)<br>Beliefs about medication (beliefs about medicine questionnaire (BMQ)) |

| | | | | | | | |
|---|---|---|---|---|---|---|---|
| Orlando M, Meredith LS, 2002 | Longitudinal, Consumer Assessment of Health Plans Survey, Patient provider relationship ratings, Data from the patient surveys | 697 | CIDI | . | US | primary care visitors in managed care organizations | Satisfaction<br>Relationship<br>Quality of care indicators |
| Pollock K, Grime J, 2002 | Follow-up, Interview | 62 | Diagnosis by GP, Depression Alliance | . | UK | GP | Entitlement to time |
| Manning C, Marr J, 2003 | Cross-sectional, Survey | 1010 | Depression Alliance | . | UK | GP | Lifestyle choices<br>Reason for stopping medication<br>Experience of recurring depression |
| Gabbay et al., 2003 | RCT, Questionnaire | 464 | Diagnosis by GP | . | UK | GP | Problem formulation<br>Agreement with GP on problem |
| Solberg et al., 2003 | Follow-up, Questionnaire, Shortform functional status survey | 274 | Chart diagnosis | . | US | PCC | Self-efficacy<br>Adherence<br>Satisfaction<br>Experience of treatment |
| Gask et al., 2003 | Cross-sectional, Interviews | 27 | Diagnosis by GP | . | UK | GP | Adherence<br>Quality of care<br>Experience of treatment |
| Wills CE, Holmes-Rovner M, 2003 | Cross-sectional, Satisfaction With Decision Scale, Decision Conflict Scale, Physician Participatory Decision-Making Style Scale, Questionnaire | 97 | Antidepressant prescription for depression | . | US | PCP | Decision-making<br>Satisfaction with decision<br>Knowledge |
| Parker et al., 2003 | Cross-sectional | 638 | 18-item questionnaire | . | Australia | GP | Attribution style |

| | | | | | | | |
|---|---|---|---|---|---|---|---|
| Templeton et al., 2003 | Longitudinal,<br>Interviews,<br>Focus groups | 20 | EPDS | Minority ethnic women | UK | GP | Experience of treatment |
| Corrigan et al., 2003 | Cross-sectional,<br>Survey | 230 | GDS-screen | Older adults | US | PCP | Discuss depression |
| Lin et al., 2003 | RCT,<br>Follow-up,<br>Interview,<br>Questionnaire | 386 | Antidepressant prescription for depression | . | US | PCC | Self-management<br>Adherence<br>Attitude to medication<br>Confidence in managing side effects |
| Van Voorhees et al., 2003 | Cross-sectional,<br>Questionnaire | 881 | CIDI | . | US | PCP | Health care utilization<br>Acceptance of care |
| Yeung et al., 2004 | Cross-sectional,<br>Explanatory Model of Interview Catalogue | 40 | CBDI | Chinese Americans | US | PCC | Chief complaint<br>Help seeking<br>Perceived causes<br>Stigma |
| Ford et al., 2004 | Case-control-matching,<br>Medical record data | 300 | Chart diagnosis | . | US | HC | Health care utilization |
| Yamada et al., 2004 | Cross-sectional | 162 | MINI | . | Japan | PCC | Complaints<br>Consultation frequency |
| Shvartzman et al., 2005 | Cross-sectional,<br>Interview | 2507 | MINI | . | Israel | PCC | Health care utilization |
| Brook et al., 2005 | RCT,<br>Patient medication records,<br>Electronic pill container | 135 | Antidepressant prescription for depression | . | The Netherlands | GP | Adherence |
| Pyne et al., 2005 | RCT,<br>Interview | 211 | IDD/ DSM IV | . | US | PCP | Acceptability of medication |
| Al-Windi et al., 2005 | Cross-sectional | 1055 | Göteborg quality of life instrument | . | Sweden | GP | Visits to GP |
| Ruoff G, 2005 | Follow-up,<br>Patient charts | 103 | Diagnosis by GP | . | US | FP | Discontinuation of medication |

| | | | | | | | |
|---|---|---|---|---|---|---|---|
| Brown et al., 2005 | Cross-sectional,<br>Observational,<br>Beliefs about Medicines Questionnaire | 192 | Antidepressant prescription for depression | . | US | FP | Beliefs about medication |
| Löwe et al., 2006 | Cross-sectional,<br>Interview | 178 | SCID /DSM IV | . | D | internal medicine outpatien t clinic | Self-management<br>Preference |
| Bogner et al., 2006 | Two-stage sampling,<br>Pill counts | 228 | SCID /DSM IV | Older adults | US | PCP | Adherence |
| Rhodes et al., 2006 | Cross-sectional,<br>Survey | 36984 | CIDI/ DSM IV | . | Canada | primary medical care | Health service use |
| Lawrence et al., 2006 | Qualitative interviews | 110 | HADS | Older adults | UK | PCP | Concept of depression |
| Givens et al., 2006 | Qualitative interviews | 42 | CES-D/ HDRS | Older adults | US | PCP | Attitude to medication |
| Gum et al., 2006 | RCT,<br>Interview | 1602 | SCID /DSM IV | Older adults | US | PCC | Preference |
| Menchetti et al., 2006 | Cross-sectional,<br>Two-phase,<br>Clinical charts | 1854 | WHO-Checklist for ICD-10 | Elderly patients | Italy | PCP | Attendance frequency |
| Akerblad et al., 2006 | RCT,<br>Sertraline-<br>/Desmethylsertraline-<br>Level,<br>Self-report,<br>Scheduled visits | 940 | Prescription of antidepressants for depression | . | Sweden | GP | Adherence |
| Backenstrass et al., 2006 | Cross-sectional,<br>Questionnaire | 607 | PHQ-9 | . | D | GP | Preference |
| Finucane A, Mercer SW, 2006 | Longitudinal,<br>Interview | 13 | BDI-II, ICD-10 chart diagnosis | . | UK | GP | Preconceptions about treatment<br>Expectations<br>Acceptability<br>Experience of treatment |

| | | | | | | | |
|---|---|---|---|---|---|---|---|
| Clever et al., 2006 | Prospective cohort,<br>Cross-sectional,<br>Rating the involvement | 1607 | CIDI/ DSM IV | . | US | PCP | Involvement in decision-making |
| Schneider et al., 2006 | Cross-sectional,<br>Fragebogen zur Erhebung von Kontrollüberzeugung zu Krankheit und Gesundheit,<br>Autonomy Preference Index | 234 | HADS | . | D | GP | Health locus of control<br>Preference for involvement |
| Mellor et al., 2006 | Cross-sectional,<br>Interview,<br>Depression Treatment Satisfaction Questionnaire | 31 | SCID | ≥65 years old | Australia | aged-care facility | Experience of care<br>Awareness of care<br>Relationship<br>Satisfaction |
| Eilat-Tsanani et al., 2006 | Follow-up,<br>Interview | 527 | EPDS | Women postpartum | Israel | FP | Consultation pattern |
| MsCracken et al., 2006 | Two-phase interview,<br>Client Service Receipt Inventory | 427 | BDI, ICD-10 | . | Ireland<br>Finland<br>Norway<br>Spain<br>UK | PCP | Service use |
| von Knorring et al., 2006 | RCT,<br>Sertraline-/Desmethylsertraline-Level,<br>Self-report,<br>Scheduled visits | 1031 | Diagnosis by GP (DSM- IV) | . | Sweden | GP | Adherence |
| Johnson et al., 2006 | Cross-sectional,<br>Question | 1140 | CIDI/ DSM IV/ chart diagnosis | . | US | PCC | Preference |
| Badger F, Nolan P, 2006 | Cross-sectional,<br>Semi-structured questionnaire | 60 | Diagnosis by GP | . | UK | PCC | Medication concordance<br>Beliefs about medication |

| | | | | | | | |
|---|---|---|---|---|---|---|---|
| | | | | | | | Role of relationship<br>Experience of medication |
| Lawrence et al., 2006 | Qualitative interviews | 110 | HADS | Older adults | UK | PCP | Help seeking |
| Mohr et al., 2006 | Cross-sectional,<br>Perceived Barriers to Psychotherapy Scale | 290 | PHQ >10 | . | US | PCC | Perceived barriers to psychotherapy |
| Brook et al., 2006 | Prospective cohort,<br>Follow-up,<br>Electronic pill container,<br>Drug Attitude Inventory | 119 | Prescription for tricyclic antidepressant for depression | . | The Netherlands | GP | Adherence<br>Drug attitude |
| Saver et al., 2007 | Cross-sectional,<br>Interview | 15 | Diagnosis by GP | . | US | PCC | Understanding of depression<br>Sources of information<br>Participation in treatment decisions<br>Barriers to treatment and information |
| Loh et al., 2007 | Follow-up,<br>Patient participation scale,<br>Scale regarding adherence | 207 | Diagnosis by GP | . | D | GP | Treatment adherence<br>Participation in treatment decisions |
| Saur et al., 2007 | Cross-sectional | 105 | SCID | Older adults | US | Academic group practice | Satisfaction with care |
| Sleath et al., 2007 | Observational,<br>Interview audiotapes | 40 | Prescribed antidepressants for depression | Veterans | US | general medicine outpatient clinic | Ask and talk about antidepressants |
| Badger F, Nolan P, 2007 | Cross-sectional,<br>Interview | 60 | Diagnosis by GP | . | UK | GP | Experience of treatment<br>Attributing recovery |
| Pollock K, 2007 | Cross-sectional,<br>Interview | 62 | Diagnosis by GP | . | UK | GP | Maintaining face in medical consultations |

| | | | | | | | |
|---|---|---|---|---|---|---|---|
| Stecker et al., 2007 | Cross-sectional | 29 | Chart diagnosis | . | US | Family medical center | Filling prescription<br>Initiation of psychotherapy<br>Stigma (The Stigma Questionnaire)<br>Beliefs about psychotherapy (The Beliefs about Psychotherapy Scale) |
| Badger F, Nolan P, 2007 | Cross-sectional, Interview | 60 | Diagnosis by GP | . | UK | GP | Self-chosen therapies |
| Weich, 2007 | Cross-sectional, Interview, Attitudes to depression and it's treatment questionnaire | 866 | CIDI/ ICD-10 | . | UK | GP | Adherence<br>Attitudes to depression and its treatment |
| Cornford et al., 2007 | Cross-sectional, Interview | 23 | HADS | . | UK | GP | Beliefs about depression |
| Johnston et al., 2007 | Cross-sectional, Interview | 111 | Diagnosis by GP | . | UK | GP | Goals for the management of depression<br>Acceptance of diagnosis |
| Dobscha et al., 2007 | RCT, Questionnaire | 314 | PHQ | Veterans | US | VC | Preference |
| Leydon et al., 2007 | Qualitative interviews | 17 | Prescription of antidepressants for depression | . | UK | GP | Barriers and facilitators to discontinuation of antidepressants |
| Ferrari et al., 2008 | Case-control study, Database | 50 | SCID, DSM IV | . | Italy | PCC | Attendance frequency |
| Druss et al., 2008 | Cross-sectional, Survey | 30801 | CIDI-SF, DSM-III-R | . | US | PC | Access to primary care |
| Frémont et al., 2008 | Cross-sectional, survey | 598 | Diagnosis by GP | . | F | GP | Causal attribution<br>Satisfaction<br>Therapeutic alliance |

| | | | | | | | |
|---|---|---|---|---|---|---|---|
| | | | | | | | Acceptance of diagnosis and treatment<br>Experience of treatment |
| Ghods et al., 2008 | Cross-sectional | 108 | Medical Outcomes Study SF-12 | African Americans and Whites | US | Group practices and communit y health centers | Communication behaviors |
| Glaesmer et al., 2008 | Cross-sectional,<br>Amount of doctor and hospital visits,<br>Scale for the Assess–ment of Illness Behavior | 1185 | PHQ-9 | ≥50 years of age | D | GP | Health service use<br>Illness behavior |
| Bazargan et al., 2008 | Cross-sectional,<br>Survey | 415 | PHQ-9 | African American and hispanic patients | US | PCC | Use of complementary and alternative medicine |
| Dernovsek et al., 2008 | Cross-sectional,<br>Questionnaire | 391 | Prescription of antidepressants for depression | . | Slovenia | public health center | Compliance |
| Damush et al., 2008 | Cross-sectional,<br>Questionnaire | 500 | PHQ-9 ≥10 | Musculoskeletal pain | US | PCC | Self-management |
| Dowrick et al., 2008 | Cross-sectional,<br>Interview | 100 | CES-D≥16 | . | UK | GP | Resilience |
| Bogner et al., 2008 | Cross-sectional,<br>Interview | 33 | CES-D | ≥65 years of age | US | PCP | Illness model |
| Russell J, Kazantzis, 2008 | Cross-sectional,<br>Medication Adherence Report Scale,<br>Beliefs about Medication Questionnaire | 85 | Diagnosis by GP | . | NZ | GP | Adherence<br>Beliefs about medication |

| | | | | | | | |
|---|---|---|---|---|---|---|---|
| Turner et al., 2008 | Cross-sectional, Interview | 27 | EPDS | Women | UK | GP | Views on medication Preference |
| Wittkampf et al., 2008 | Cross-sectional, Interview | 17 | PHQ, SCID, HDRS | . | The Netherlands | GP | Views on screening and treatment Acceptance of diagnosis |
| Chew-Graham et al., 2009 | RCT, Interview | 28 | Prescription of antidepressants for depression | Women postpartum | UK | GP | Disclosure of symptoms Understanding postnatal depression Barriers of disclosure |
| Bogner et al., 2009 | Qualitative interviews | 102 | CES-D | 65 and older | US | PCP | Views regarding antidepressant medication |
| van Geffen et al., 2009 | Retrospective, Contact file, Pharmacy dispensing registration database | 965 | Prescription of antidepressant for depression | . | The Netherlands | GP | Contact with GP Initiation of antidepressant treatment and filing prescriptions |
| Hérique A, Kahn JP, 2009 | Retrospective, Cross-sectional, Questionnaire | 632 | Diagnosis by GP | . | F | GP | Compliance to antidepressants |
| Wittink et al., 2009 | Cross-sectional, Interview | 47 | CES-D | Older African Americans | US | PCP | A Faith-Based Explanatory Model of Depression |
| Beattie et al., 2009 | Longitudinal, Interview | 24 | Diagnosis by GP | . | UK | GP | Completion of online-CBT Expectations of online CBT Experience of online CBT |
| Raue et al., 2009 | Longitudinal, Randomized, Care manager records, Participants' reports, Questionnaire | 60 | SCID | . | US | PCP | Adherence Initiation Expectation Preference |
| Menchetti et al., 2009 | Two-phase, PCP records, Patient chart | 250 | WHO ICD-10 Symptom Checklist for Depression | . | Italy | PCP | Visiting frequency Presenting complaints |
| Suija et al., 2009 | Cross-sectional, Registration forms | 1094 | CIDI | . | Estonia | FP | Visits to FD |

| | | | | | | | |
|---|---|---|---|---|---|---|---|
| Danielsson et al., 2009 | Cross-sectional, Interview | 20 | ICD-10, chart diagnosis | . | Sweden | health care center | Source and course of depression |
| Hodges et al., 2009 | Cross-sectional, Questionnaire | 100 | Diagnosis by GP | Cancer patients | UK | GP | Preference |
| Prins et al., 2009 | Cross-sectional, Perceived Need for Care Questionnaire | 622 | CIDI | . | The Netherlands | primary care center | Perceived need for care |
| Vega et al., 2010 | Cross-sectional, Stigma Checklist | 200 | PHQ-9 score ≥10 | Latino patients | US | PCC | Stigma |
| Dickinson et al., 2010 | Cross-sectional, Interview | 36 | Antidepressant prescription for depression | ≥ 75 years of age | UK | GP | Understanding depression<br>Views on long term use of antidepressants<br>Barriers to discontinuation |
| Malpass et al., 2010 | Longitudinal, Interview | 10 | PHQ-9, diagnosis by GP | . | UK | GP | Patients' experiences of using the PHQ-9 in primary care consultations |
| Barg et al., 2010 | Follow-up (randomized), Interview | 24 | CES-D | Older patients | UK | PCC | Experience of treatment |
| Hansson et al., 2010 | Cross-sectional, Open-ended question | 303 | Diagnosis by GP | . | Sweden | health care center | Belief about the cause of depression |
| van Geffen et al., 2010 | Follow-up, Observational, Data from dispensing records, Questionnaire | 110 | Antidepressant prescription for depression | . | The Netherlands | GP | Initiation and continuation of antidepressant treatment<br>Health beliefs |
| Palmer et al., 2010 | Longitudinal, Interview | 474 | CESD ≥ 16 | . | Australia | GP | Desires towards depression treatment |
| Newman et al., 2010 | Cross-sectional, Interview | 40 | PHQ-9 | Gay men | Australia | GP | Ascribed role of the GP |

| | | | | | | | |
|---|---|---|---|---|---|---|---|
| Gask et al., 2011 | Cross-sectional, Interview | 15 | Diagnosis by GP | Pakistani women | UK | GP | Understanding persistence of depression<br>Views on depression treatment |
| Mergl et al., 2011 | RCT, Fragebogen zur Messung der Psychotherapiemotivation (Item 36) | 145 | CIDI, DSM IV | . | D | PCP | Attitude to psychotherapy preference |
| Malpass et al., 2011 | Longitudinal, Interview, Consultation records | 10 | Antidepressant prescription for depression | . | UK | GP | Unvoiced agenda |
| Saito et al., 2011 | Cross-sectional, Interview | 5 | Diagnosis by GP | . | Japan | PCP | Perceived triggers for depression<br>Opinions on treatment |
| Fortney et al., 2011 | Cross-sectional | 395 | PHQ 9 | Veterans | US | VA medical center | Adherence<br>Acceptability of antidepressants<br>Health beliefs (The Depression Health Beliefs Inventory)<br>Reasons for non-adherence |
| Callister et al., 2011 | Qualitative interviews | 20 | Postpartum Depression Screening Scale, Spanish Version | Hispanic women postpartum | US | Community health center | Help seeking |
| Körner et al., 2011 | Cross-sectional, Interview | 40 | PHQ 9 | Gay men | Australia | GP | Understanding of depression<br>Experience of depression |
| Prins et al., 2011 | Follow-up, Perceived Need for Care Questionnaire | 568 | CIDI, DSM IV | . | The Netherlands | GP | Perceived need for care |
| Deen et al., 2011 | RCT, Depression Health Beliefs Inventory, One Item regarding acceptability, | 360 | PHQ9 ≥12 | . | US | PCP | Health beliefs<br>Satisfaction with care<br>Acceptability of treatment<br>Patient-centeredness of care |

| | | | | | | | |
|---|---|---|---|---|---|---|---|
| | One scale regarding satisfaction,<br>Experience of Care and Health Outcomes Survey | | | | | | |
| Lynch et al., 2011 | Cross-sectional,<br>Beliefs about Depression Questionnaire | 334 | Diagnosis by GP | . | UK | GP | Beliefs about depression |
| Bell et al., 2011 | Follow-up,<br>Perceived Barriers Index | 1054 | PHQ-9 | . | US | PCP | Barriers of disclosure |
| Elwy et al., 2011 | Cross-sectional,<br>Interview | 30 | PHQ-9 ≥11 | . | US | PCP | Illness perception |
| van den Boogaard et al., 2011 | Cross-sectional,<br>Medical records,<br>Causal attributions inventory | 120 | CIDI | . | The Netherlands | GP | Visits to GP<br>Causal attribution |
| Gensichen et al., 2012 | Cross-sectional,<br>Interview | 41 | Diagnosis by GP | . | D | GP | Perspective on collaborative care |
| Kwong et al., 2012 | Cross-sectional,<br>Stigma assessment scale,<br>Interview | 42 | PHQ-9 | Chinese American | US | community health center | Help seeking<br>Perceived cause of depression<br>Perceived stigma<br>Reasons that prevent individuals from participating in a depression treatment program |
| Chew-Graham et al., 2012 | Cross-sectional,<br>Interview | 19 | Diagnosis by GP | Older people | UK | GP | Accessing help<br>Reasons to not present with depression |
| Hansson et al., 2012 | Follow-up,<br>Questionnaire | 184 | Diagnosis by GP, HADS | . | Sweden | primary health care center | Perceived improvement factors |
| Calderón Gómez et al., 2012 | Observational,<br>Discussion groups, | 31 | Diagnosis by GP | . | Spain | FP | Views on treatment |

| | | | | | | | |
|---|---|---|---|---|---|---|---|
| | Interview | | | | | | |
| Cornford et al., 2012 | Cross-sectional,<br>Interview,<br>Focus group | 24 | Diagnosis by GP | Opioid dependency | UK | GP | Causes of depression<br>Experience of depression |
| Hansen et al., 2012 | Cross-sectional,<br>Focus group | 19 | PHQ-9 | Low-income Latinos with diabetes | US | PCC | Help seeking<br>Barriers to help seeking |
| Dwight Johnson et al., 2013 | Cross-sectional,<br>Survey | 63 | SCID | Older mexican men | US | PCC | Preference for treatment |
| Simmonds et al., 2013 | Cross-sectional,<br>Interview | 30 | PHQ2 | Patients with coronary heart disease | UK | GP | Links between CHD and depression<br>Experience of CHD and depression |
| Busch et al., 2013 | Cross-sectional,<br>Survey | 2890 | EURO-D-Skala ≥4 | ≥50 years of age | D | GP | Health service utilization |
| Bennett et al., 2013 | Cross-sectional,<br>Interview | 41 | CIDI | Chronic depression | UK | GP and practice nurses | Experience of care<br>Treatment initiation<br>Adherence<br>Barriers to care |
| Kales et al., 2013 | Observational,<br>Follow-up,<br>Brief Medication Questionnaire | 188 | Antidepressant prescription for depression | Old age | US | PCC | Adherence |
| Izquierdo et al., 2014 | Two-stage,<br>Interview | 47 | CIDI | Older Latinos | US | PCC | Experience of visits to PC for mental health |
| Keeley et al., 2014 | Cross-sectional,<br>Interview | 30 | Diagnosis by PCP,<br>ICD-9 | . | US | PCP | Conceptualization of depression<br>Expectations of depression care<br>Experience of depression care |
| DeJesus et al., 2014 | Cross-sectional,<br>Questionnaire | 125 | Diagnosis by GP | . | US | primary care site | Self-management strategies<br>Satisfaction with CCM for depression<br>Opinions of care managers |

| | | | | | | | |
|---|---|---|---|---|---|---|---|
| | | | | | | | Attitudes towards care management |
| Magnezi et al., 2014 | Cross-sectional, Patient Activation Measure | 278 | PHQ-9 | . | Israel | PCC | Patient activation |
| Solberg et al., 2014 | Cross-sectional, Questionnaire | 1168 | Diagnosis by GP, PHQ-9 ≥7 | . | US | PCC | Shared decision-making |
| Osei-Bonsu et al., 2014 | Observational, Brief COPE, Interview | 23 | PHQ-9 | Veterans | US | VC | Coping strategies |
| Serrano et al., 2014 | Observational, Longitudinal, Simplified Medication Adherence Questionnaire, Drug Attitude Inventory | 29 | DSMIV, HADS ≥17 | . | Spain | primary care center | Adherence<br>Attitude to drugs |
| Fosgerau CF, Davidsen AS, 2014 | Observational, Conversation analysis | 28 | Diagnosis by GP, ICD-10 | . | Denmark | GP | Patients' perspectives on antidepressant treatment |
| Edwards et al., 2014 | Cross-sectional, Questionnaire | 1478 | Diagnosis by GP | . | UK | GP | Satisfaction with telehealth use<br>Advantages of telehealth use |
| Jaffray et al., 2014 | Cross-sectional, Interview | 29 | Diagnosis by GP | . | UK | GP | Attitude to antidepressants |
| Waltz et al., 2014 | Cross-sectional, Questionnaire | 761 | PHQ-9 | Veterans | US | VC | Satisfaction<br>Treatment preference |
| Holloway et al., 2015 | Longitudinal, Open-ended question | 57 | PHQ-9 | Vision-impaired | Australia | GP | Uptake of GP referral |
| Chen et al., 2015 | RCT, Explanatory Model Interview Catalogue | 190 | PHQ-9 ≥10 | Chinese American | US | community health center | Chief complaint<br>Name of problem/ perceived cause of depression<br>Experience of help sought |
| Knowles et al., 2015 | Cross-sectional, Interview | 31 | PHQ-9 ≥10 | Physical long-term condition | UK | PCP | Experience of collaborative care |
| Lynch et al., 2015 | Longitudinal, | 227 | Diagnosis by GP | . | UK | GP | Beliefs about depression |

| | | | | | | | |
|---|---|---|---|---|---|---|---|
| | Beliefs about Depression Questionnaire | | | | | | |
| Lee King et al., 2015 | Cross-sectional,<br>Treatment Beliefs Scale,<br>Medication Beliefs Scale,<br>Agnew Relationship Measure,<br>Goldring Patient–Provider Scale,<br>Interview | 198, 29 | Chart diagnosis | . | US | PCP | Treatment beliefs<br>Relationship<br>Experience of depression |
| Horevitz et al., 2015 | Two-phase,<br>Chart review,<br>Interview | 431, 16 | Chart diagnosis, PHQ-9 | Latinos | US | health care center | Taking up referral to behavioral health<br>Relationship<br>Factors influencing decision to take up referral |
| Knowles et al., 2015 | Cross-sectional,<br>Interview | 36 | PHQ-9 ≥10 | . | UK | collaborative care | Experience of computerized CBT |
| Patel et al., 2015 | Case-control study | 142 | SCID | . | UK | GP and practice nurses | Attendance |
| Larsen et al., 2016 | Longitudinal,<br>National register | 1332 | HADS-D ≥ 8 | Heart disease | Denmark | GP | Contact with GP |
| von Faber et al., 2016 | Qualitative interviews | 38 | Geriatric Depression Scale (GDS-15) | 75 and older | The Netherlands | GP | Coping strategies |
| Elwy et al., 2016 | Prospective observational study | 271 | PHQ-9 | Veterans | US | VA PCC | Illness perceptions (the illness perceprions questionnaire - revised (IPQ-R)) |
| Campbell et al., 2016 | Cross-sectional,<br>Questionnaire | 761 | PHQ-9 | Veterans | US | VC | Care engagement<br>Stigma<br>Agreement with depression label<br>Treatment preference |

| | | | | | | | |
|---|---|---|---|---|---|---|---|
| | | | | | | | Report of provider behavior<br>Received treatment |
| Keller et al., 2016 | Cross-sectional, Interview | 24 | PHQ-8, history of depression | Women | US | PCP | Experience of disclosure<br>Barriers and facilitators to disclosure |
| Vuorilehto et al., 2016 | Longitudinal, Monitoring records, Questionnaire | 100 | SCID/ DSM IV | . | Finland | GP | Visits to GP<br>Adherence to treatment<br>Initiation of treatment |
| Chen et al., 2016 | Follow-up, Explanatory Model Interview Catalogue | 168 | CB-PHQ-9 ≥10 | Chinese immigrants | US | health care center | Stigma |
| Magnani et al., 2016 | Cross-sectional, Question | 170 | HDRS ≥13 | . | Italy | PCP | Preference |
| Davis et al., 2016 | Follow-up, Questionnaire | 761 | PHQ-9 ≥10 | Veterans | US | VA collaborative care | Satisfaction with care<br>Preference<br>Experience of PC-related mental health care |
| Rossom et al., 2016 | Observational, Question regarding care quality, Modified Patient Assessment of Chronic Illness Care | 792 | PHQ-9 | . | US | PCC | Care rating<br>Experience of patient-centered care |
| Sternke et al., 2016 | Focus groups data | 18 | PHQ-9 ≥10 | Chronic pain | US | VC and PCC | Experience of empathy |
| Wikberg et al., 2016 | Recorded focus groups | 9 | Diagnosis by GP | . | Sweden | GP | Patients' experience of the use of MADRS-S in primary care |
| Moise et al., 2017 | Cross-sectional, Morisky Medication Adherence Scale, Control Preference Scale | 195 | PHQ-8 ≥10 | Hypertension | US | PCC | Adherence<br>Preference for decision-making |

| | | | | | | | |
|---|---|---|---|---|---|---|---|
| Robinson et al., 2017 | Longitudinal,<br>Interview | 29 | Diagnosis by GP | . | UK | PCP | Explanations for discrepancies between changes in PHQ-9 and GRC |
| Saint Aurnalt et al., 2017 | Cross-sectional | 209 | CES-D | Japanese immigrants | US | PC | Illness interpretation |
| Green et al., 2017 | Cross-sectional,<br>Interview,<br>Focus groups | 28, 12 | PHQ-9 ≥10 | Latinos | US | collaborative care | Adherence to antidepressants<br>Receiving information about antidepressants<br>Experience of taking medication |
| Gunn et al., 2018 | Longitudinal,<br>Survey | 789 | CES-D ≥10 | . | Australia | GP | Health service use<br>Self help |
| Stark et al., 2018 | Cross-sectional,<br>Interview | 12 | Geriatric Depression Scale (GDS) ≥6 | Older patients | D | GP | Attitudes concerning depression<br>Views on depression treatment |
| Lopez et al., 2018 | Cross-sectional,<br>Stigma Concerns About Mental Health Care Measure,<br>Social Distance Measure,<br>Latino Scale for Antidepressant Stigma Measure,<br>Depression Knowledge Measure | 319 | PHQ-9 | Hispanic women | US | PCC | Stigma<br>Knowledge on depression |
| Dorow et al., 2018 | Cross-sectional | 641 | Diagnosis by GP | . | D | PCP | Preference for treatment |
| Taylor et al., 2018 | Cross-sectional,<br>Interview | 13 | MINI | Older patients | UK | collaborative care | Views on collaborative care for depression |
| Waitzfelder et al., 2018 | Observational,<br>Health record data | 241251 | Diagnosis by GP, ICD-9 | . | US | primary care site | Initiation of treatment |
| Jones et al., 2018 | Cross-sectional,<br>Survey | 27362 | ICD-9, chart diagnosis | . | US | VC | Number of clinic visits<br>Experience of care |
| Vannoy et al., 2018 | Cross-sectional,<br>Observational, | 77 | SCID | Older men | US | PCP | Attitudes towards discussing suicide |

| | | | | | | | |
|---|---|---|---|---|---|---|---|
| | Interview | | | | | | Patients' perspectives on what primary care providers can do to help prevent late-life suicide |
| Pung et al., 2018 | Cross-sectional, Interview | 16 | PHQ-2 | . | Australia | PCP | Patterns of mobile app use among patients with depressive symptoms |
| Avey et al., 2018 | Cross-sectional, Questionnaire | 125 | PHQ-9 ≥10 | Alaskan Native, American Indian | US | PCC | Preference |
| Gordon et al., 2018 | Cross-sectional, Interview | 16 | Diagnosis by GP | Older people | UK | GP | Understanding of depression |
| Carmassi et al., 2018 | Cross-sectional | 75 | SCID | . | Italy | GP | Attendance<br>Empowerment (Empowerment in patients with Affective and Schizophrenic Disorders) |
| Canty et al., 2019 | Cross-sectional, Interview | 17 | EPDS | Women postpartum | US | PCC | Barriers and facilitators to mental health care postpartum<br>Experience with depressive symptoms |
| Aznar-Lou et al., 2019 | Cross-sectional,<br>Beliefs About Medicines Questionnaire,<br>The Brief Illness Perception Questionnaire | 263 | Diagnosis by GP, SCID | . | Spain | GP | Beliefs about illness and medicine |
| Schomerus et al., 2019 | Longitudinal,<br>Self-Identification as Having Mental Illness Scale,<br>Social Distance Scale,<br>Questionnaire | 188 | PHQ-9 ≥8 | . | D | GP | Help seeking intention<br>Self-identification as having a mental illness<br>Prejudice and discrimination<br>Literacy for depression<br>Perceived need for care |
| Liu et al., 2019 | Cross-sectional, Medical records | 2976 | PHQ-8 | Older patients with osteoarthritis | US | group health cooperati ve | Health care use |

| | | | | | | | |
|---|---|---|---|---|---|---|---|
| Aragonès et al., 2019 | RCT, Questionnaire, Medical records | 274 | SCID | Musculoskeletal pain | Spain | primary care center | Health care resource use<br>Adherence to program<br>Satisfaction |
| Richards et al., 2019 | Cross-sectional, Interview | 37 | PHQ-2 | . | US | primary care center | Experience of being asked about suicidality |
| Backhaus et al., 2020 | Cross-sectional, Question | 641 | Diagnosis by GP, PHQ-9 ≥ 5 | . | D | GP | Treatment preference |
| Pitrou et al., 2020 | Cross-sectional | 1624 | Questionnaire based on DSM-IV | 65 years and older | Canada | PCP | Satisfaction with care |
| Pirard et al., 2020 | Cross-sectional, Questionnaire | 454 | HADS-D≥8 | Survivors of terrorist attacks | F | GP | Mental health support use |
| Kloppe et al., 2020 | Cross-sectional, Interview | 26 | PHQ-9 | 60 years and older | D | GP | Relief seeking style |
| Gomà-I-Freixanet et al., 2020 | Case-control-matching, Medical records | 238 | Goldberg Anxiety–Depression Scale ≥2 | . | Spain | PCP | Self-initiated consultations |
| Heinz et al., 2021 | Cross-sectional, Questionnaire, Depression Stigma Scale | 430 | Depression screening questionnaire, ICD 10 | . | D | GP | Reported complaints<br>Stigma |
| Cornwell et al., 2021 | Cross-sectional | 92535 | PHQ-2 | Veterans | US | integrated primary care | Initiation of treatment |
| Poole et al., 2022 | Cross-sectional, Interview | 41 | Diagnosis by GP | Older patients, Long-term conditions | UK | GP | Experience of depression |
| Gartland et al., 2022 | Prospective cohort study | 1507 | CES-D | Mothers | Australia | GP | Visits to GP<br>Trust in PCP (Health Care Relationship Trust (HCRT) Scale<br>Willingness to seek help |
| Grung et al., 2022 | Cross-sectional, Survey | 250 | Self-identification as depressed | . | Norway | GP | Experience of care |

| | | | | | | | |
|---|---|---|---|---|---|---|---|
| Af Winklerfelt Hammarberg et al., 2022 | Cluster randomized controlled trial | 376 | Diagnosis by GP | . | Sweden | PCC | Self-efficacy<br>Preferences |
| Taylor et al., 2022 | Cross-sectional, Interview | 14 | PHQ-2 | . | UK | GP | Self-help strategies<br>Confidence in own ability |
| Harrison et al., 2023 | Feasibility study | 18 | PHQ-9 | . | UK | GP | Adherence to treatment |
| Jeffery et al., 2023 | Cohort study | 73808 | Chart diagnosis | Patients with type 2 diabetes | UK | PCP | Discontinuation of antidepressants |

Appendix 3: List of abbreviations

| Abbreviation | Full phrase |
|---|---|
| BDI | Beck Depression Inventory |
| CBDI | Chinese translation of the Beck Depression Inventory |
| CB-PHQ-9 | Chinese Bilingual version of the 9-item Patient Health Questionnaire |
| CBT | Cognitive Behavioral Therapy |
| CES-D | Center for Epidemiologic Studies – Depression Scale |
| CIDI | Composite International Diagnostic Interview |
| CIDI-SF | Composite International Diagnostic Interview – Short Form |
| CPG | Clinical practice guidelines |
| DIS | Diagnostic Interview Schedule |
| DSI-SS | Depressive Symptom Inventory – Suicidality Subscale |
| DSM | Diagnostic and Statistical Manual of Mental Disorders |
| EPDS | Edinburgh Postnatal Depression Scale |
| FD | Family doctor |
| FP | Family practitioner |
| GDS | Geriatric Depression Scale |
| GHQ-12 | 12-item General Health Questionnaire |
| GP | General practitioner |
| GRC | Global Rating of Change |
| HADS | Hospital Anxiety and Depression Scale |
| HAMD | Hamilton Rating Scale for Depression |
| HDRS | Hamilton Depression Rating Scale |
| HRSD | Hamilton Rating Scale for Depression |
| HSCL-90 | Hopkins Symptom Checklist |
| ICD | International Statistical Classification of Diseases and Related Health Problems |
| IDD | Inventory to Diagnose Depression |
| MINI | Mini-International Neuropsychiatric Interview |
| N/A | Not applicable |
| OECD | Organisation for Economic Co-operation and Development |
| PC | Primary care |
| PCP | Primary care professionals |
| PHQ | Patient Health Questionnaire |
| RCT | Randomized controlled trial |
| RQ | Research question |
| SCID | Structured Clinical Interview for DSM-IV |
| Short-CARE | Short Comprehensive Assessment and Referral Evaluation |
| WHO | World Health Organization |
